# Variant-risk-exon interplay impacts circadian rhythm and dopamine signaling pathway in severe psychiatric disorders

**DOI:** 10.1101/2022.08.09.22278128

**Authors:** Karolina Worf, Natalie Matosin, Nathalie Gerstner, Anna S. Fröhlich, Anna C. Koller, Franziska Degenhardt, Holger Thiele, Marcella Rietschel, Madhara Udawela, Elizabeth Scarr, Brian Dean, Fabian J. Theis, Janine Knauer-Arloth, Nikola S. Mueller

## Abstract

In psychiatric disorders, common and rare genetic variants cause widespread dysfunction of cells and their interactions, especially in the prefrontal cortex, giving rise to psychiatric symptoms. To better understand these processes, we traced the effects of common and rare genetics, and cumulative disease risk scores, to their molecular footprints in human cortical single-cell types. We demonstrated that examining gene expression at single-exon resolution is crucial for understanding the cortical dysregulation associated with diagnosis and genetic risk derived from common variants. We then used disease risk scores to identify a core set of genes that serve as a footprint of common and rare variants in the cortex. Pathways enriched in these genes included dopamine regulation, circadian entrainment, and hormone regulation. Single-nuclei-RNA-sequencing pinpointed these enriched genes to excitatory cortical neurons. This study highlights the importance of studying sub-gene-level genetic architecture to classify psychiatric disorders based on biology rather than symptomatology, to identify novel targets for treatment development.

## INTRODUCTION

Major psychiatric disorders, including schizophrenia, major depressive disorder (MDD), and bipolar disorder (BD), are among the top ten leading causes of burden globally ^1^. These disorders share common pathophysiological, clinical and biological characteristics ^2–4^, and a key feature of their pathology is altered connectivity and synaptic plasticity in prefrontal cortex (PFC) neural circuits. These brain areas play a crucial role in cognition and executive functioning, processes which are commonly impacted across the spectrum of psychiatric disorders ^5^. Acknowledging these shared features, cross-disorder psychiatric studies enable the examination of patients based on their common biological processes, rather than solely based on phenotypic features, which constitutes a priority research approach in psychiatry ^4,6–8^. While alterations in the living human brain can only be studied with neuroimaging methods, postmortem specimens provide unique resolution to examine disease-related changes in neural circuits at the cellular level and transcriptome-wide molecular processes.

Genome-wide association studies (GWAS) are an effective tool to investigate the genetic predisposition of complex psychiatric disorders. In recent years, large schizophrenia, BD and MDD GWAS have identified hundreds of genetic loci associated with these disorders from hundreds of thousands of samples at the level of single variants. For example, Trubetskoy et al. ^9^ performed a GWAS on schizophrenia with 76,755 cases and 243,649 controls and identified 287 genetic risk loci that were strongly enriched in gene expression of the Brodmann area 9 (BA9) PFC region. Based on the single variant GWAS results, a new approach to calculate polygenic risk scores (PRSs) was established ^10^. The PRS for an individual is based on their cumulative genetic risk to be affected by a disorder. In psychiatric research, PRSs still need improvement before they may be used as predictive diagnosis tools; for example, the PRS for schizophrenia explained only 7.3% of the variance in genetic liability ^9^. With improved PRS methods and larger sample sizes in GWAS, PRSs can be refined to provide insight into the genetic architecture of psychiatric disorders, and computer-aided diagnosis of psychiatric disorders could become a trend in the future ^11,12^.

In parallel to the GWAS approach studying common single nucleotide polymorphisms (SNPs), examination of rare or de novo, highly penetrant variants may also clarify the genetic factors that contribute to psychiatric disorders. For example, Gulsuner et al. ^13^ showed that schizophrenia de novo mutations imply a disruption of neurogenesis and transcription. Wainschtein et al. ^14^ hypothesised that the lower heritability estimate of common SNPs compared to population-based estimates is due to rare variants, which are often located in regions of low linkage disequilibrium (LD) and are more likely protein altering. This is also consistent with previous observations that rarer and evolutionarily younger SNPs exhibit higher SNP heritability in many complex traits including BD, MDD, and schizophrenia ^15^.

Complementary studies are aiming to pinpoint molecular changes in the brain on the level of gene expression. This can be achieved using postmortem brain samples, well established for large cohorts like Genotype-Tissue Expression (GTEx) ^16^, CommonMind Consortium (CMC) ^17,18^ and PsychENCODE ^19^. More recent developments in single-cell/nucleus RNA-sequencing (sc/snRNA-seq) technologies have enabled the study of cell-type-specific gene expression ^20,21^. The first single-cell atlas for schizophrenia was recently published and identified differentially expressed genes mainly in inhibitory and excitatory neurons ^22^.

The majority of GWAS significant variants are located in non-coding regions of the genome, thus not directly affecting the protein sequence ^23,24^. However, some of these variants might function by influencing gene expression, also called expression quantitative trait loci (eQTL) ^25^. Previous studies have shown that SNPs with evidence of association with schizophrenia are more likely to be eQTLs ^26^. The majority of these studies focus on eQTL analysis at the gene-level ^27–29^, with some recently looking at the sub-gene-level, such as SNPs associated with differential splicing (splicingQTL) ^16,30,31^. Alternative splicing affects genes with multiple exons and provides a mechanism by which genes can produce diverse ranges of gene products. Alternative splicing effects have been previously reported for schizophrenia ^32–36^. The first comprehensive study of brain splicingQTLs found evidence for a strong enrichment among schizophrenia GWAS loci ^34^. Since most of the known disease-causing mutations occur in exons, genetic methods that focus on protein-coding regions in the genome are valuable for identifying potentially pathogenic mutations ^37^. Despite this, no study has investigated genetic effects on exon expression in psychopathology using a targeted approach (e.g. arrays specifically designed to accurately quantify exons vs RNA sequencing). The accurate quantification of exons is essential to understand the genetic architecture of schizophrenia beyond the gene-level ^38,39^.

Understanding genetic effects on exons, and tracing this to the affected cell types, represents a key gap in knowledge. In this study, we performed a multi-factorial investigation that dissects the effects of disease-status, common variants and polygenic risk scores on gene expression in the human prefrontal cortex (BA9) of cross-diagnostic psychiatric subjects. We demonstrate that examination of gene expression at the exon-level is essential to fully capture molecular changes. In parallel, we complement the genotype-/risk-modulated exon-level differential expression with rare and damaging variants. By examining both common genetic effects on exon expression and rare variant damaged genes, we found that pathways involved in cortisol and neurotransmitter regulation, specific to excitatory neurons of cortical layers of BA6, 10 and 11, are enriched in severe psychiatric disorders. Our results highlight how studying sub-gene-level genetic architecture can elucidate new knowledge regarding the development of severe psychiatric disorders and assist in classifying these disorders based on biology rather than symptomatology ^40^. It also highlights how novel cellular targets for improved drug development can be precisely identified.

## RESULTS

### Cortical gene expression at the resolution of exons associates with diagnosis of psychiatric disorders

Gene expression analyses are typically performed at the level of gene models, which may level out effects of higher granularity ultimately resulting in alternative splicing. Especially in highly complex diseases, subtle but globally molecular changes may occur at this lower resolution, such as differentially expressed exons. To address this potential drawback of gene model analysis in the context of highly complex psychiatric cross-disorder, we profiled expression of postmortem dorsolateral prefrontal cortex (DLPFC) tissue of BA9 from 169 individuals aged 18-87 years (68 schizophrenia, 24 BD and 15 MDD and 62 controls, Table 1) using Affymetrix exon microarrays. We designed an exon array data pre-processing, analysis and testing strategy that allowed us to compare differential expression results across gene, transcript and exon levels (Figure S1). 6.5 million array probes were summarised into 242,443 exons, 100,750 transcripts and 17,447 gene models, while only one exon or transcript per gene model was subjected to multiple testing correction, so as not to confound the correction of genes with hundreds of exons (Figure S1 and methods). Using linear regression modelling for differential expression analysis, we included age, cause of death (CoD), postmortem interval (PMI) and genetic ancestry covariates (Dim1-Dim4, Table 1 and Figure S2) as covariates to account for effects of clinical and demographic variables. The BA9 expression data was complemented by matching genotype data for all individuals, and following quality control and imputation, 9,572,139 common variants were yielded.

**Table 1:** Summary of the human postmortem brain samples (Dataset 1) and their demographic and clinical variability. For continuous data the mean ± standard error and for categorical data the categories separated by dashes are given for control and case subjects as well as the different diagnosis types (schizophrenia, BD, MDD) of the cases. Age = years, postmortem interval (PMI) = hours (h), sex = = males (M) and females (F), suicide = yes (Y) and no (N) and cause of death (CoD) = grouped into natural (N), non-violent (NV) and violent (V) death. Dim 1 – 4 = the first four dimensions of the ancestry information calculated with the sample genotypes.

| Diagnosis | N | Age (year) | Sex (M/F) | PMI (h) | pH | RIN | Suicide (Y/N) | CoD (N/NV/V) | Dim 1 | Dim 2 | Dim 3 | Dim 4 |
| --- | --- | --- | --- | --- | --- | --- | --- | --- | --- | --- | --- | --- |
| Controls | 62 | 49.33<br>± 2.0 | 49/13 | 41.89<br>± 1.9 | 6.37<br>± 0.03 | 7.69<br>± 0.1 | 0/62 | 56/1/5 | -8e-5<br>± 0.002 | -0.0008<br>± 0.001 | -0.0019 ± 0.002 | 0.0009<br>± 0.002 |
| Cases | 107 | 50.06<br>±1.7 | 74/33 | 42<br>± 1.3 | 6.36<br>± 0.02 | 7.6<br>± 0.08 | 48/59 | 39/24/44 | -0.0005<br>± 0.002 | -0.0011<br>± 0.001 | -9e-5<br>± 0.001 | -0.0005<br>± 0.001 |
| SCZ | 68 | 46.88<br>± 2.1 | 54/14 | 43.59<br>± 1.6 | 6.31<br>± 0.03 | 7.82<br>± 0.1 | 33/35 | 30/10/28 | 0.0002<br>± 0.002 | -0.0016<br>± 0.002 | -0.0008<br>± 0.002 | -0.0004<br>± 0.001 |
| BIP | 15 | 57.61<br>± 3.6 | 8/7 | 39.57<br>± 3.9 | 6.27<br>± 0.04 | 7.07<br>± 0.1 | 5/10 | 6/4/5 | 0.0013<br>± 0.005 | 0.0005<br>± 0.003 | 0.0032<br>± 0.003 | -0.0038<br>± 0.004 |
| MDD | 24 | 54.35<br>± 3.7 | 12/12 | 38.98<br>± 2.9 | 6.54<br>± 0.04 | 7.42<br>± 0.1 | 21/3 | 3/10/11 | -0.003<br>± 0.002 | -0.0008<br>± 0.003 | -6e-5<br>± 0.003 | 0.0009<br>± 0.002 |

To increase statistical power and cut across traditional nosological boundaries, samples with schizophrenia, MDD and BD were grouped to one cross-disorder diagnosis, with schizophrenia being the dominant diagnosis. We found that zero, six and 2,223 genes (FDR<10%) respectively at the level of gene models, transcripts and exons, respectively, were associated with cross-disorder cases in BA9 of human DLPFC (Figure 1A-D and Figure S3A-C). Tissue enrichment analysis with FUMA ^41^ confirmed that genes of the differentially expressed exons were particularly enriched for GTEx v8 brain regions (Figure S3D). KEGG pathway analysis of exon-level hits identified six pathways involved in cell-cell interactions, cell motility and various organismal systems (Figure S3E). Extracellular matrix (ECM)-receptor interaction (FDR=7×10^−5^) and complement and coagulation cascade (FDR=0.027) pathways contained 69% and 91% up-regulated genes, respectively. Conversely, 65% of the genes in the axon guidance (FDR=0.046) pathway were down-regulated (Figure S3E).

**Figure 1:**
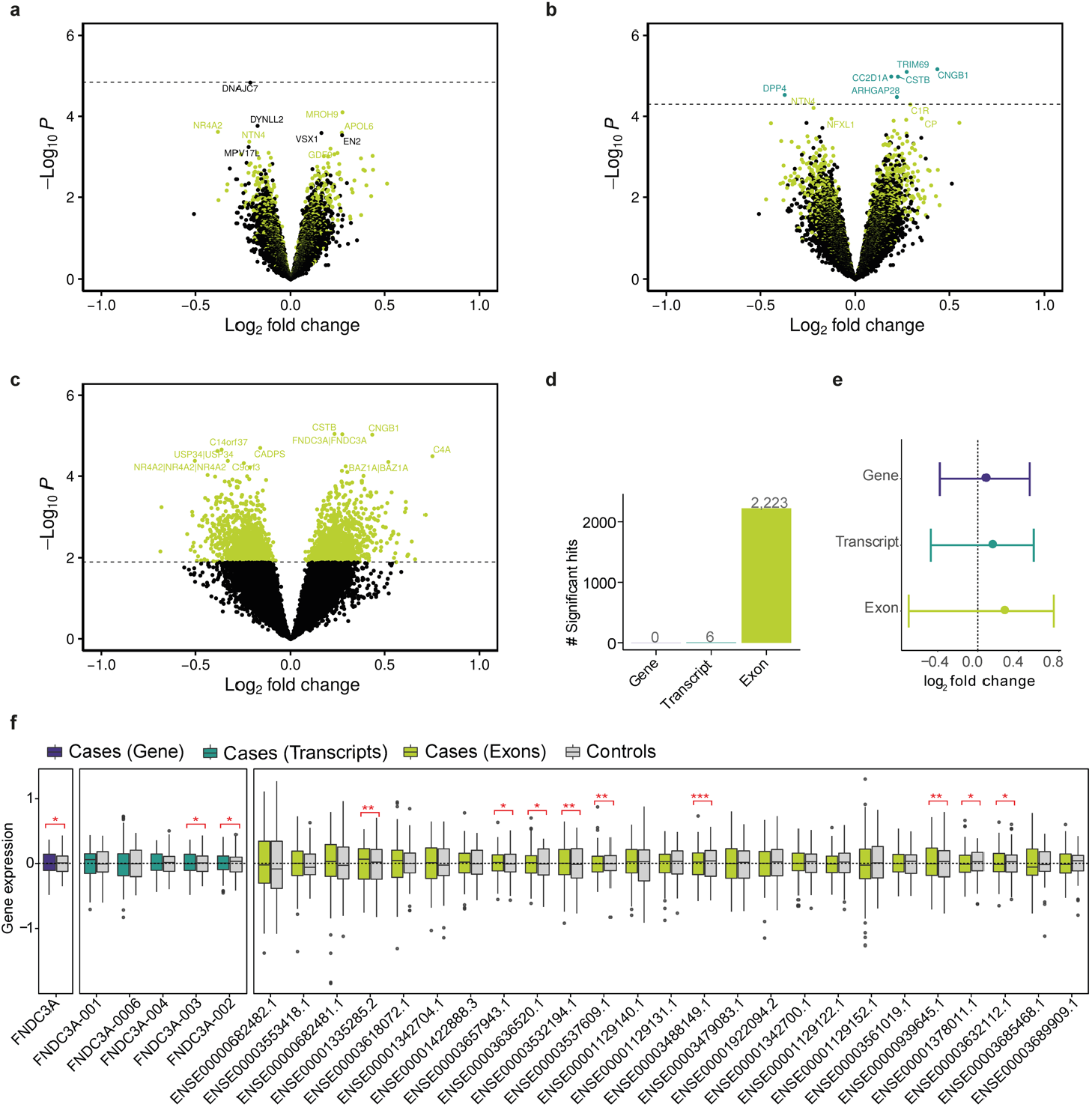
Differential expression results. Volcano plots of the (a) gene-, (b) transcript- and (c) exon-level differential expression analysis results. On the x-axis the log_2_ fold changes and on the y-axis the -log_10_(P-values) are given. The dashed horizontal line represents the significance threshold (FDR < 0.1) with all significant hits colored in turquoise for transcript-level and light green for exon-level differentially expressed gene hits. 51% of the exon-level (1,139 of 2,223 genes) and 83% transcript-level differentially expressed genes (5 of 6 genes) are upregulated. (d) Bar plot of significant differentially expressed genes for gene-, transcript- and exon-level. (e) Forest plot showing the log_2_FC range and median of the absolute log_2_FC (dot) of the 2,223 exon-level differentially expressed genes for all three levels. The 2,223 genes exhibit a larger magnitude of changes on the exon-level (median absolute log_2_FC = 0.23, range of -0.69 to 0.75) compared to the two other levels (transcript median absolute log_2_FC = 0.11, range of -0.47 to 0.55, and gene median absolute log_2_FC = 0.047, range of -0.38 to 0.51). (f) Boxplots show the effect of cross-disorder diagnosis on *FNDC3A* expression, separately for each level: full gene, five of its six transcripts and 25 of its 46 exons with available expression values for cases (purple, turquoise or light green) and control subjects (grey). The x axis indicates expression residuals. Transcripts and exons were sorted by the difference in median expression from cases to controls. Asterisks represent the significance level for the corresponding p-values.

As anticipated for the cohort size, gene-level analysis did not identify any differentially expressed genes. The six genes differentially expressed at the transcript-level were also differentially expressed at the exon-level. To better understand the gene-, transcript-, and exon-level difference, we analysed the fold changes (FCs) of all exon hits at the respective transcript- and gene-level and observed a 2.1- and 4.9-fold decrease in the median of the absolute log_2_FCs, respectively (Figure 1E). Next, we selected the *fibronectin type III domain containing 3A* (*FNDC3A*), an ECM-glycoprotein that plays vital roles during tissue repair, as a multi-exon gene regulated only at the exon-level ^42^. Only one of 26 exons was significantly associated with diagnosis (ENSE00003488149, FDR=0.0539), while most exons showed no expression differences (Figure 1F). This suggests that the median summarization of many probes per gene compensates for the pronounced regulation of the few sub-gene alterations.

In addition, we explored the level difference for other covariates to understand if sub-gene resolution implies an information gain for other factors (Figure S3A,G and Table S2 to Table S6). Genetic ancestry, captured by one ancestry dimension (Dim2) of the genotype data, showed prominent differences at the gene-level. Age-related effects exerted similar impacts at each level (Figure S3F). Notably, age and pH strongly influenced gene expression. These findings together suggest that exon-level differences are related to diagnosis, rather than a global effect of brain expression.

### Common variants associated with changes in cortical gene expression are enriched for psychiatric cross-disorder GWAS traits

In parallel to the association of cross-disorder diagnosis labeling alone with DLPFC expression, we set out to determine the common variants that influence this expression independently of diagnosis. To that end, we computed *cis*-eQTLs on all three levels, yielding 44,040 full gene-eQTLs, 203,069 transcript-eQTLs and 477,352 exon-eQTLs at FDR<5% (Figure 2A, Figure S4 and Table S8 to Table S10). The number of uniquely identified SNPs and genes increased from gene-, transcript-to exon-eQTL analysis (Figure 2B). In contrast, 6% genes (n=72 out of 1,114) of the gene-eQTL were unique to this level, while 65% genes (n=4,133 out of 6,401) were exclusively detected at the exon-eQTL level (Figure 2B). To illustrate the exon-level-specific effect, we used the *calcium-binding gene neurocalcin delta* (*NCALD*) with SNP rs505460, where only one exon out of 13 exons (ENSE00001231633, FDR = 4×10^−45^) was differentially expressed in a variant-dependent manner (Figure 2C). Notably, 49% of the differentially expressed exon-level genes (n=1,090 out of 2,223 genes) were in common with exon-eQTL genes.

**Figure 2:**
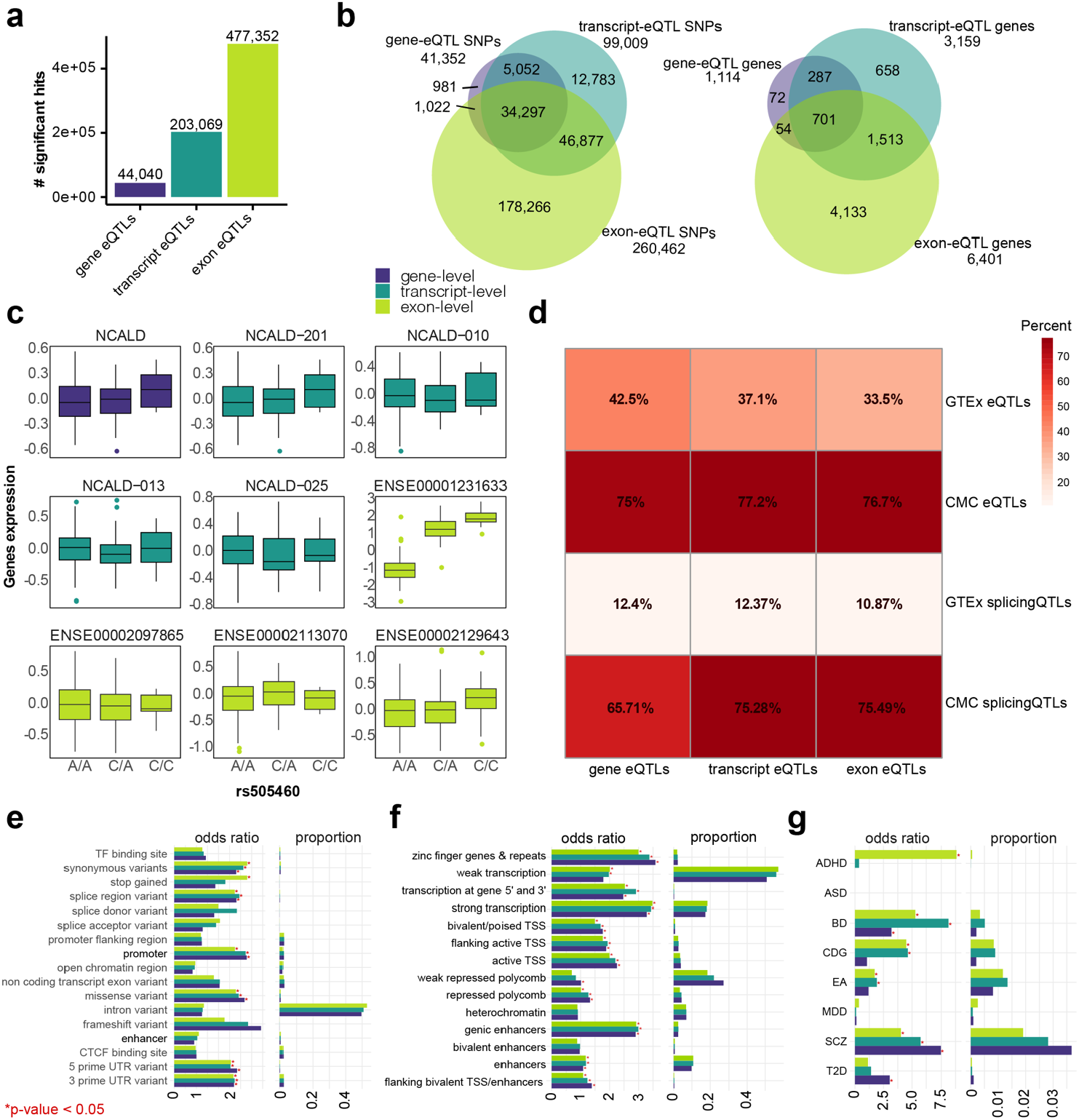
eQTL results and downstream their genomic features enrichment analysis. (a) Bar plot of significant eQTLs for gene, transcript and exon-level. (b) Venn diagrams visualising the eSNPs and eQTL genes overlap all three levels. (c) An example association plot for rs505460-NCALD expression on all three levels. We selected the full gene, four out of 30 transcripts and four out of 68 exons. The SNP shows only the effect on the expression of exon ENSE00001231633. The x axis indicates expression residuals. (d) Overlap between genes from the three eQTL levels and eQTL and splicingQTL genes from GTEx or CommonMind Consortium datasets in the human cortex. (e) Genomic overlap between variant effect predictor (VEP). (f) 15-state model of the Roadmap Epigenomics Project measured in DLPFC. (g) GWAS traits and eSNPs from all three levels. Results are presented as bar plots showing the odds ratios with Asterisk indicating significant p-values for the enrichment test, and proportions of overlap with the original Datasets. ADHD attention-deficit/hyperactivity disorder, ASD autism spectrum disorder, BD bipolar disorder, CDG cross disorder meta-analysis, EA educational attainment, MDD major depressive disorder schizophrenia schizophrenia and T2D type 2 diabetes.

In order to evaluate the robustness of our eQTLs, we compared the eQTL genes, to previously reported PFC (BA9) eQTLs from GTEx v8 (425 donors) and the CMC (467 donors with schizophrenia and BD; see Figure 2D). Overall, overlap of the general cohort with GTEx data was expectedly lower than to the disease-matched CMC cohort. The largest overlap >75% was yielded for our exon-eQTLs and CMC eQTLs/splicingQTLs (Figure 2D), indicating reliable identification of prefrontal cortex eQTLs in our study when compared to CMC data.

To better understand the properties of our *cis*-eQTLs, we characterized the eQTL (e)SNPs according to genomic locations and regulatory features. Most eSNPs across all three levels were localised within intronic regions (Figure 2E) and were significantly enriched for synonymous variants, splice regions, promoters, missense, 5’ and 3’ UTR variants. Interestingly, eSNPs of exon-eQTLs were specifically enriched for stop gains. Integrative analysis of chromatin states of the DLPFC using ChromHMM ^43^ mainly showed that eSNPs of all three levels are associated to both active and repressive chromatin marks (Figure 2F). The eSNPs of gene-eQTLs showed a specific enrichment to weak repressed polycomb elements (Figure 2F).

Next, we explored how the identified eSNPs map onto genetic risk for psychiatric disorders. To do this, we tested whether eSNPs were overrepresented amongst variants associated with psychiatric disorders from large-scale GWAS of the Psychiatric Genomics Consortium (PGC) ^44^ and non-psychiatric phenotypes as negative controls (Figure 2G). We detected significant enrichment of eSNPs on all three levels at a nominal GWAS p-value cut-off associated with schizophrenia and BD, Figure 2G. For transcript- and exon-level eSNPs, we found significant enrichment in cross-disorder (CDG) and educational attainment (EA) GWAS. Only exon-level eSNPs were enriched for attention deficit hyperactivity disorder (ADHD). In summary, the gene-, transcript- and exon-eQTL hits may be significantly attributed to the genetic architecture of psychiatric diseases, with exon-level data specifically capturing the largest overlap (Table S12).

### Multi-SNP effects are associated with cross-diagnostic disease risk influence cortical exon expression

We next followed up on the exciting link between exons differentially regulated in BA9 of psychiatric subjects, and their encoding on genomic loci that were previously identified in large psychiatric GWAS. We specifically sought to determine how to effectively encode genetic disease risk in a variable, to associate it again with exon expression in the prefrontal cortex. To that end, we implemented PRS analysis, which allows us to compress the full genetic risk load to a cumulative sum of risk variants per individual. We calculated the risk for the three major psychiatric disorders, namely schizophrenia, MDD and BD, and the cross-disorder GWAS from PGC for all 169 samples. Notably, the PRSs did not correlate with the available covariates (Figure S5). Further, we focused on exon-level expression as it provided the strongest signal in our cohort. Using the identical analysis approach as for differential gene expression analysis, we computed the exon expression quantitative trait (eQT)-Score using the individual PRS scores of each GWAS together with exon expression (Figure 3A).

**Figure 3:**
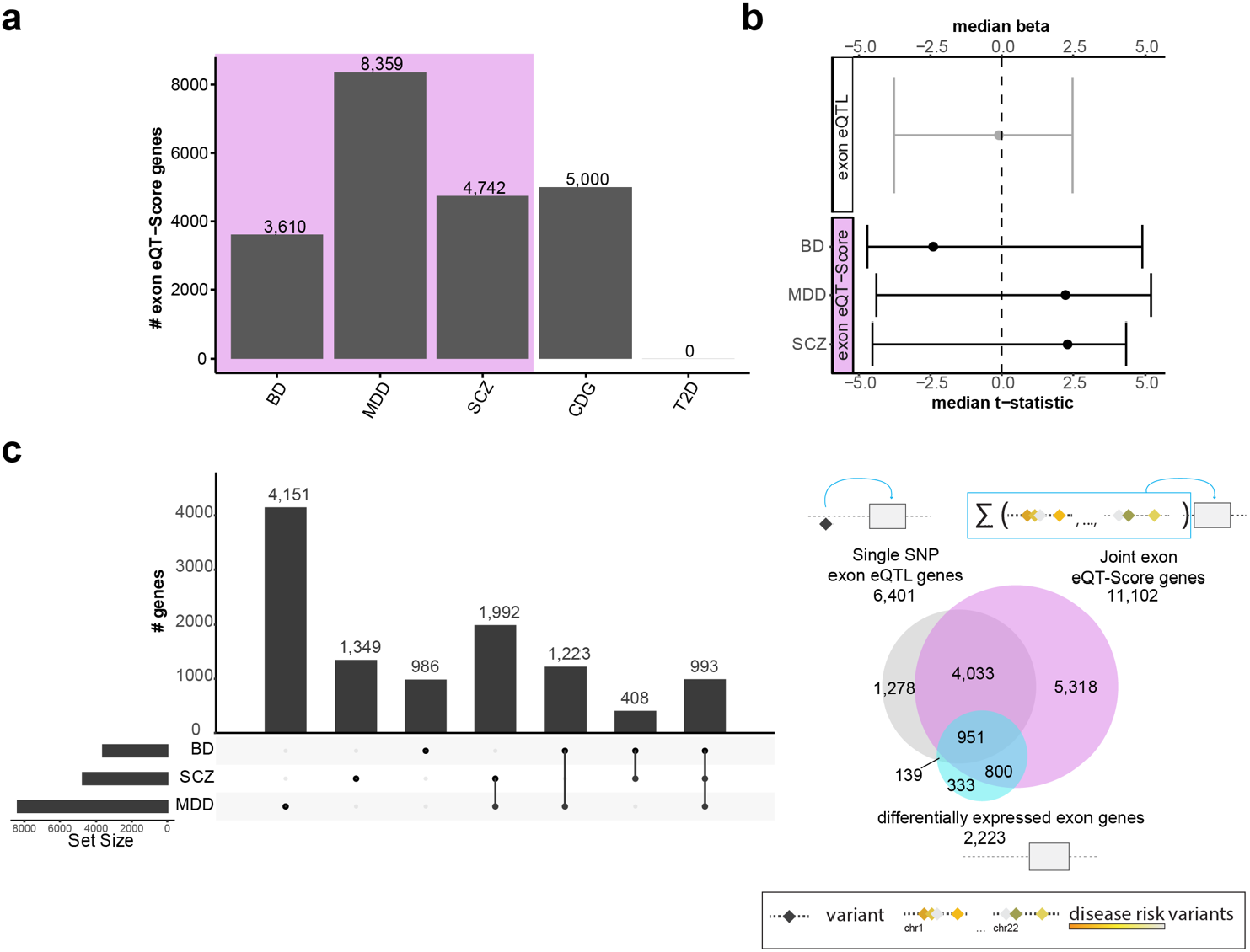
Multi-SNP effects and their influence on cortical expression. (a) Bar plot shows the significant number of exon expression -polygenic risk score associations, also called exon expression quantitative trait score (exon eQT-Score). The y-axis shows the counts of exon eQT-Score genes and the x-axis displays the GWAS in the eQT-Score calculation. (b) Forest plot of the effect size of the exon eQTL genes and exon eQT-Score genes. The y-axis shows median beta or t-statistic and the x-axis displays the GWAS used in the eQT-Score calculation. Exon eQT-Score genes exhibit an overall larger effect size (median absolute schizophrenia t-stats = 2.55, range from -4.53 to 4.32, median absolute MDD t-stats = 2.46, range from -4.39 to 5.2, median absolute BD t-stats = 2.63, range from -4.81 to 4.89) compared with the minimal effects for single exon eQTL genes (median absolute beta = -0.1, range from -3.77 to 2.48, p-value Wilcoxon test < 2.2 × 10^−16^). (c) Upset plot shows the overlap between ADHD, BD, MDD and schizophrenia GWAS exon eQT-Score genes. Each row corresponds to a GWAS study and black dots indicate an intersection. Bar chart on the top indicates the size of the intersection. Bar chart on the left shows the number of exon eQT-Score genes. (d) Venn diagram shows the overlap between the exon eQTL genes (grey), the joint genes of the ADHD, BD, MDD and schizophrenia GWAS exon eQT-Scores (purple) and the differentially expressed exon genes (blue). SCZ schizophrenia, MDD major depressive disorder, BD bipolar disorder, CDG cross disorder meta-analysis and T2D type 2 diabetes.

The exon eQT-Score analysis identified multiple genes for schizophrenia, MDD and CDG (Figure 3A), while the absolute number of associated genes (3,610-8,359, corresponding to a total of 11,102 unique genes) is larger than for the cross-disorder diagnosis (2,223, c.f. Figure 1B). This suggests that the full load of the genetic disease architecture provides more information than the binary diagnosis. As negative control GWAS we used type 2 diabetes (T2D), which yielded zero hits.

Most exon eQT-Score inherent genes are in common between MDD and schizophrenia (n=2.290 with 63% shared effect direction), followed by MDD and BD (n=2,217, with 44% shared effect direction), schizophrenia and BD (n=825 with 59% shared effect direction), and schizophrenia, MDD and BD (n=995 with 32% shared effect direction; Figure 3C). The union of exon eQT-Scores of the three single GWAS (BD, MDD and schizophrenia; n=11,102 genes and 16,199 exons, Table S14) hereafter is referred to as joint exon eQT-Score set.

The joint exon eQT-Score set covered 78.8% of the exon-level genes associated with cross-disorder diagnosis (Figure 3D), thus a large fraction of regulated genes is attributable to genetic disease risk. Asking whether the multi-SNP approach of the exon eQT-Score set provides more information, we compared the set to single SNP eQTL results. In total, 77.9% of the single-SNP exon-eQTLs overlap with the multi-SNP joint exon eQT-Score set. The effect sizes of the multi-SNP approach were overall larger than for single-SNP (Figure 3B).

We further investigated if the identified joint exon eQT-Scores were also differentially expressed for diagnosis (Table S1iii, 15.8% and n=1,751 DE genes) and show single SNP-effects in *cis* (Table S10, 44.9% and n=4,985 exon eQTL genes), see Figure 3D.

### Rare and common variants share risk for cross-diagnostic psychiatric disorders

Since common variants only account for 23% of heritability in schizophrenia ^45^, we used the recent results from the Schizophrenia Exome Meta-Analysis (SCHEMA) Consortium, which included 24,248 cases and 97,322 controls, and identified 244 genes with rare variants associated with schizophrenia (Dataset 2, see Methods) ^46^. We examined how genes might be affected by rare and common variants by comparing common single *cis*-eSNP effects, common multi-SNP effects and rare harmful effects. We identified a core set of 110 genes present in all three data types (see Figure 4A). We performed pathway enrichment analysis of these 110 core set genes to identify if they share the same function. This analysis indicated that one-fifth of the genes (n=42, 38.2%) were significantly enriched in eight KEGG pathways including brain-related pathways such as cortisol synthesis and secretion (FDR=0.026) and dopaminergic synapse (FDR=0.038), indicating a similar functional impact of rare and common variants in psychiatric disorders (see Figure 4B).

**Figure 4:**
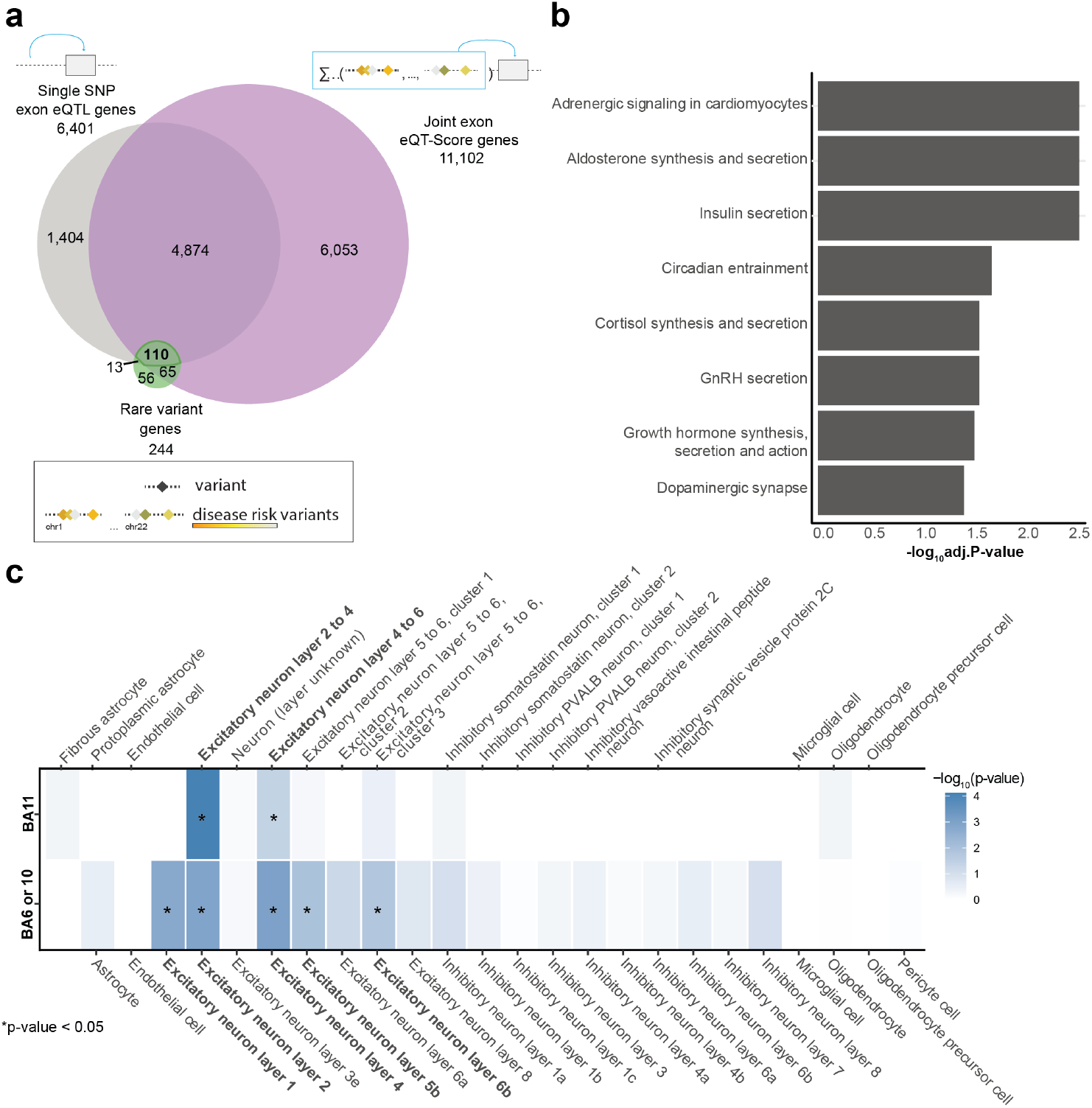
Genes xdisrupted by rare and common variants. (a) Venn diagram of the overlap between single SNP exon eQTL genes (grey), joint exon eQT-Score genes (purple) and genes altered by rare variants in SCHEMA consortium data (green). (b) Bar plot shows the KEGG pathway enrichment for the core set of genes (n=110). Only significant gene sets are shown. The y-axis shows the enriched KEGG pathways/gene sets and the x-axis shows the -log_10_ of the adjusted p-value (FDR). (c) Heatmap depicting cell type specificity for the core gene set of enrichment signals defined at gene-level from snRNA-seq data (BA11: Dataset 2 and B6 or B10 ^48^). The star indicates significant enrichment p-value.

To characterise the cell-type specificity of these core genes, we used snRNA-seq data from the human orbitofrontal cortex (BA11, Data set 3, Table 2, see Methods). Among the 18 delineated cell-type clusters, the core genes were substantially enriched in excitatory neurons (layer 2-4: p=7.67×10^−5^ and layer 4-6: p=0.04, Figure 4C). We replicated the enrichment of our core genes in excitatory neurons with Web-based Cell-type Specific Enrichment Analysis of Genes (WebCSEA) ^47^ in snRNA-seq data from BA6 (premotor) and BA10 (frontal pole) cortices ^48^ (p-values<0.016, Figure 4C). We next aimed to gain deeper mechanistic insight into the subset of core genes found significantly enriched in BA11 excitatory neuron layer 2-4 or 4-6 (p<0.05 and absolute fold change >1.15, n=29 genes). An overview of all annotated categories of these genes is given in Figure 5A. The majority (83-90%) of these genes overlapped with previously identified cortical gene-level eQTL genes and isoform-level eQTL genes (CMC Dataset), 38-53% of the exon-eSNPs were enriched in weak transcribed regions or enhancers, and 86% of the genes were significantly associated with PRS for MDD, followed by 66% for CDG and 59% for schizophrenia. Only seven (24%) of the genes overlapped with the differentially expressed exon-level genes. The genes with most annotations were the *Calcium Voltage-Gated Channel Subunit Alpha1 C* (*CACNA1C*), *Ryanodine Receptor 2* (*RYR2*,) and *Glutamate Ionotropic Receptor NMDA Type Subunit 2A* (*GRIN2A*) genes (Figure 5 and S6). *GRIN2A* showed enrichment in excitatory neurons, GWAS SNPs for schizophrenia, BD, MDD and CDG, actively-transcribed states and enhancer measured in DLPFC, synonymous variants (VEP) and has been replicated as cortex e/splicingQTL gene in the CMC dataset (Figure 5A-B). In addition, *GRIN2A* was down-regulated in cases (mean residualized expression in control=0.04 and cases=-0.02, Figure 5C) with controls showing a slightly higher CDG PRS than cases. In addition, it was down-regulated in schizophrenia and MDD exon eQT-Score genes. Alterations in the Glutamate systems are widely reported in schizophrenia ^49^, and accordingly we found significantly down-regulation *GRIN2A* exon expression (ENSE00001327113.1) for SNP rs7188841 (Figure 5D).

**Table 2:** Demographic of the two samples used for snRNA-seq (Dataset 3). Given are the diagnosis, sex differentiated between male (M) and female (F), age in years, post-mortem interval (PMI) in hours (h), cause of death (CoD) and ethnicity.

| Group | Sex (M/F) | Age (year) | PMI (h) | RIN | CoD | Ethnicity |
| --- | --- | --- | --- | --- | --- | --- |
| Controls | M | 60 | 21.5 | 7.7 | cardiac | European |
| Controls | F | 63 | 50 | 6.7 | cardiac | European |

**Figure 5:**
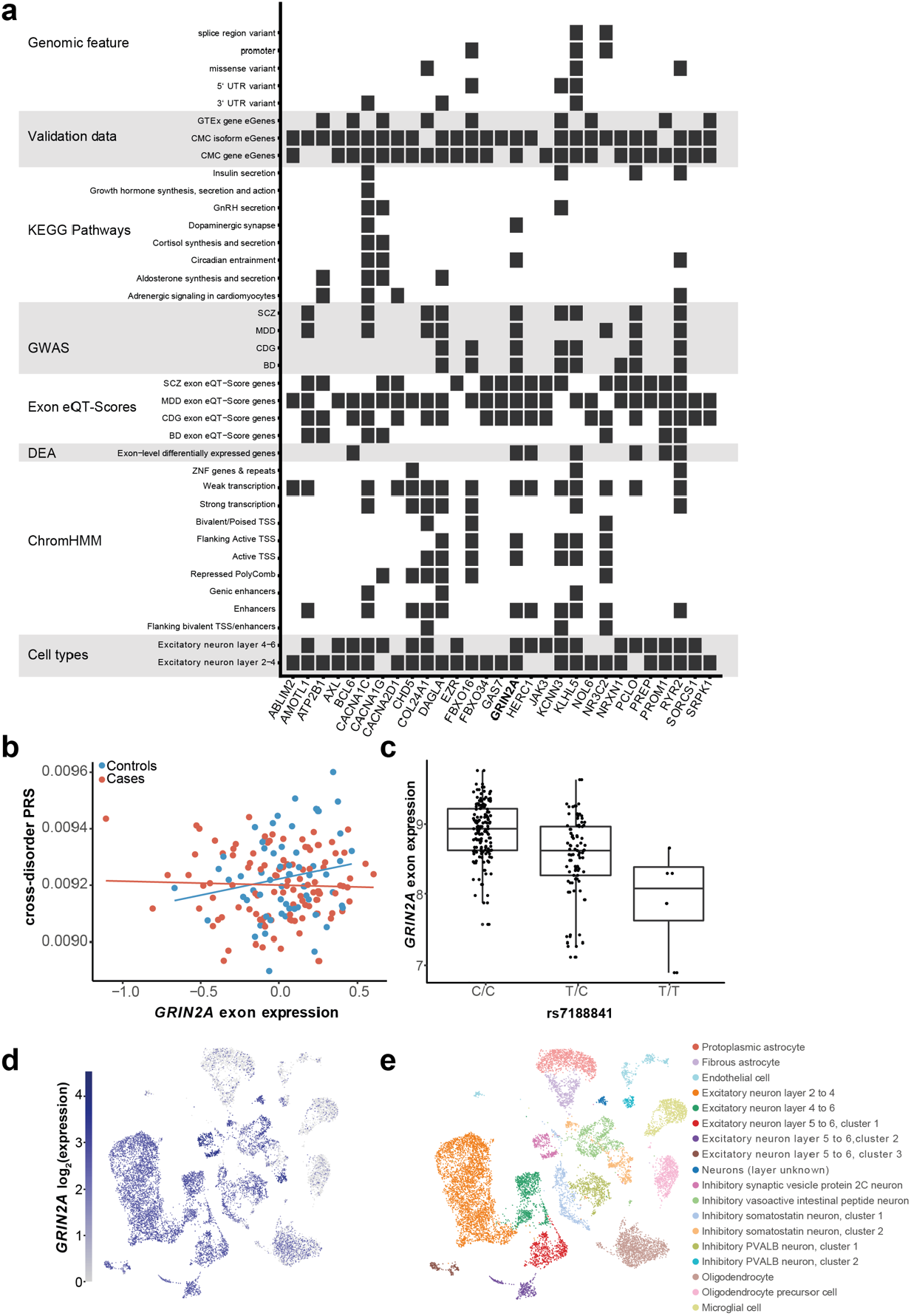
Missense gene *GRIN2A*, a core set gene enriched in excitatory neurons. (a) Heatmap of the 29 genes from the core gene set that were significantly enriched (FDR < 0.05 and absolute fold change >1.15) in at least one of the overrepresented two BA11 cell types. The plot shows in which datasets the genes are significant. (b) UMAP of the *Glutamate Ionotropic Receptor NMDA Type Subunit 2A* (*GRIN2A*) expression, where grey denotes minimal expression and blue high expression. (c) Association plot for CDG PRS-ENSE00001323909.1 exon expression of *GRIN2A*, separated into cases and controls (CDG exon eQT-Score). The x axis indicates expression residuals; the y axis shows CDG PRS values. (d) The box plot indicates the top exon-eQTL of *GRIN2A*: ENSE00001327113.1 exon expression -rs7188841. The y axis indicates expression values; the x axis shows genotypes.

## DISCUSSION

More than 95% of genes with multiple exons are alternatively spliced ^50^. Alternative splicing in the brain is highly complex compared to other tissues ^51^ and suggested to play an important role in neuronal development and function ^52^. Splicing defects have been suggested to be involved in the pathogenesis of psychiatric disorders, including schizophrenia ^53–55^. In this study, we provide the first profiling of exon-specific expression across the human cortex. Examination of exon expression is an important but relatively underexplored mechanism in psychiatry research, with most studies using methods that do not accurately quantify exons (such as RNA sequencing) or summarising expression by averaging across whole transcripts or genes. We found no significant main effects of psychiatric cross-disorder diagnosis at the gene-level and few at the transcript-level (n=6 genes). However, the exons of 2,223 genes were differentially expressed between cases and controls (see Figure 1D). This highlights how the subtle effects of exons are often overlooked but are crucial to uncover disease pathogenesis. Our findings are in line with previous work that has shown an exon-based strategy improves the detection of small but effective expression changes that might be missed by gene-based approaches ^56,57^.

Most of the transcript- and exon-level differentially expressed genes we identified are supported by other studies. For example, our top hit at the transcript-level, *Cyclic Nucleotide Gated Channel Subunit Beta* 1 (*CNGB1*, exon-level p-value: 9.2×10^−6^, transcript-level p-value: 6×10^−6^) was found to be differentially expressed in exome sequencing studies of various psychiatric disorders including schizophrenia and bipolar disorder ^58^. Our top hit at exon-level, *Cystatin B* (*CSTB*, exon-level p-value: 8.8×10^−6^, transcript-level p-value: 1×10^−5^) was also differentially expressed in a schizophrenia case-control study ^59^. *Dipeptidyl peptidase 4* (*DPP4*, exon-level p-value: 0.0017, transcript-level p-value: 2.9×10^−5^) found at both levels is also a hit in the 2014 GWAS of schizophrenia ^60^ and 19 of the 106 protein-coding genes (17.92%) found in the recent GWAS of schizophrenia ^9^ intersect with our exon-level differentially expressed genes.

By integrating the transcriptome and genetic variation data, we found that the fraction of significant eQTLs at the exon-level was two times higher than at the transcript-level and even ten times higher than at the gene-level (Figure 2A). An exon-level specific eQTL is *Neurocalcin Delta* (*NCALD)*, visualised in Figure 2D. The effect of the differentially expressed exon ENSE00001231633 was not detectable at the transcript and gene-level, likely because of the compensating effect of the other non-differentially expressed exons at this locus. *NCALD* is a brain-enriched highly conserved neuronal calcium sensor protein ^61^ and has been associated with multiple neurological disorders. In a genetic rat model of schizophrenia, *NCALD* was downregulated ^62^, and SNPs in *NCALD* have been associated with autism and BD ^63,64^. In a recent study *NCALD* has been linked to adult hippocampal neurogenesis ^65^. Interestingly, 75-77% of our significant exon-level eQTL genes overlap with previously published postmortem eQTL genes, and 65-75% with splicingQTL genes, demonstrating the validity of our results as well as the added value of examining exon-specific expression. The analysis of variant effects coming from exon-eQTLs leads to a high accumulation of gene flanking regions as well as synonymous and missense variants. Only exon eSNPs are enriched in stop-gained variants, which could imply a higher protein-damaging impact than seen on the transcript-or gene-level. Furthermore, since common variants have a very small effect size and rare variants are sought to contribute more to genetic risk of psychiatric disorders, but are rarely detected in GWAS ^66,67^, our results model the joint effects of common and rare variants. Our investigation of rare coding variants from whole exomes of 24,248 schizophrenia cases and 97,322 controls from the SCHEMA consortium detected mutations that affect the same genes as those found for exon-eQTL (50% overlap, Figure 4A), suggesting a common basis of loci for schizophrenia. The SNPs of our exon-level eQTLs were enriched for GWAS SNPs of BD, ADHD, cross-disorder, educational attainment, and schizophrenia, confirming that schizophrenia is a complex and polygenic disorder that shares many common genetic and phenotypic features with other psychiatric disorders. Despite the complexity and shared genetic basis of psychiatric diseases, we show that our results are stable and can be supported by other larger studies.

Symptoms and disease progression of patients diagnosed with psychiatric disorders are highly heterogeneous within the same disorder. In parallel, different psychiatric diseases are highly comorbid, supporting the model of a transdiagnostic phenotype. In addition, psychiatric diseases are highly polygenic and a recent study showed that GWAS and eQTL studies cover systematically different variants ^68^. With our study, we demonstrate an alternative way to integrate exon-level expression with genetic variation by using eQT-Scores. This approach provided more information than binary case-control diagnosis, with 1.6-3.8 fold more genes differentially expressed (Figure 3A,D) and 22.3-47.5 times higher effect sizes (Figure 3B) than single exon-level eQTL genes. Our findings contradict a study by Curtis et al. ^69^ who could not find any associations between PRS and gene-level gene expression in the postmortem dorsolateral prefrontal cortex (BA9/46) of schizophrenia subjects from the CommonMind Consortium. This is likely because gene-level approaches underestimate the gene expression level changes, and we gain information by estimating gene expression at certain exons using a more precise measure of exon expression. Together, our study shows that a more sensitive approach examining exon-level expression combined with polygenic continuous phenotypes (in the form of a PRS) improves the results towards risk prediction. In the future, larger sample sizes and advanced polygenic risk score calculations that include functional annotation information (including cell type or epigenetic marks) could lead to even better exon eQT-Score results and provide more precision for identifying novel diagnostic tools.

By integrating all possible genetic modelling of the transdiagnostic psychiatric phenotype (eQTL, eQT-Score and rare variants), we found a core set of 110 genes, which combined rare and common risk for psychopathology. This core set revealed a network of genes involved primarily in brain functions (dopaminergic synapse and circadian entrainment) as well as peripherally produced hormones (cortisol, insulin, aldosterone and GnRH) (Figure 4B). A deficit in dopamine, particularly in the prefrontal cortex, is the longest standing theory of schizophrenia pathogenesis, given dopamine agonists can induce psychosis and antipsychotic drugs impact on the dopaminergic signaling ^70^. Interestingly, circadian dysfunction is a frequently described phenomena in major psychiatric disorders ^71^ with a complex reciprocal relationship to dopamine, which entrains the master clock of the suprachiasmatic nuclei ^72^. An important function of the suprachiasmatic nuclei is to regulate hormone secretion ^73^, consistent with our identification of cortisol, aldosterone, insulin and GnRH regulation, hormones which all follow a circadian rhythm ^74,75^. Gene expression patterns in the prefrontal cortex are also shown to have diurnal rhythms in the CommonMind Consortium ^76^. We showed that our core gene set which implicated circadian rhythms is enriched in excitatory neuron subtypes of BA6, 10 and 11. Projections from the orbitofrontal cortex to cholecystokinin-expressing neurons of the suprachiasmatic nucleus have been previously reported ^77^ and several studies have also highlighted the impact of different cortical layer subtypes in schizophrenia ^78^, which has been linked to cognitive function ^79^. Cortical neurons also appear to be vulnerable to the effects of stress in psychopathology ^80^, relevant given the implication of cortisol in our core gene set, and is likely linked to circadian entrainment. Future studies focusing on single nucleic omics will continue to uncover salient cell types involved in psychiatric disease, such as Ruzicka et al. ^81^ who also found neuronal cell types to be the most affected in schizophrenia postmortem brains.

To demonstrate how to leverage the multi-level information from genetics (common/rare), and tissue transcriptomics (bulk exon array expression and snRNA-seq), we chose to zoom into a small set of our core genes (n=29 genes). The justification for this focus was based on the collected evidence from all our functional annotations, including the enrichment in single-cell types to explore how genetics impacts expression of specific cells in the brain, and how this is related to disease. Two standout genes were the *CACNA1C* <u>82</u>,<u>83</u> and the *Glutamate Ionotropic Receptor NMDA Type Subunit 2A* (*GRIN2A*) genes ^84^, both genes encoding related proteins that moderate neuronal excitability. *CACNA1C*, which encodes the α1C subunit of L-type voltage-gated calcium channels, was among the first GWAS hits in schizophrenia ^85^. Alterations to this calcium channel are associated with depressive-like symptoms in the prefrontal cortex ^86^, with the risk-associated SNP (rs1006737) correlated with increased cortical function during executive cognition and higher cortical *CACNA1C* mRNA levels during brain development ^87^. *GRIN2A* was recently reported to be highly associated with schizophrenia in the largest exome-sequencing study to date ^46^, and NMDA receptor hypofunction and the resultant glutamate dysregulation are among the most highly implicated and replicated finding in postmortem brain studies examining the schizophrenia cortex, thought to be intrinsically linked to dopamine neurotransmission and its dysfunction ^88,89^. The NMDA receptor is composed of multiple subunits, with the composition of this receptor switching from containing the NR2B subunit to the NR2A (*GRIN2A*) subunit over development. Alterations in this subunit switch may alter the maturation of cortical circuits and lead to NMDA receptor hypofunction ^88^. Further analysis of the core gene set could lead to new insights into schizophrenia and underpin the finding of a common basis in psychiatric disorders.

Taken together, the presented data reveals the importance of studying gene expression at the exon level directly in the human brain, and incorporating multi-level holistic datasets to identify important, combined risk gene sets with pathological relevance. This approach enabled us to identify cortical excitatory neurons as a cell-type likely to be increasingly implicated in psychopathology, as well as identifying key genes (*GRIN2A* and *CACNA1C*) which may constitute clear cellular targets for further study and treatment development for psychiatric disorders.

## METHODS

### Study samples, tissue collection and processing

#### Data set 1: Human postmortem brain samples (exon array data)

The primary cohort (Dataset 1) has been previously described in Scarr et al. ^90^ and Dean et al. ^91^. Briefly, postmortem DLPFC tissue from 169 adult subjects aged 18-87 years with either schizophrenia (n=68), MDD (n=24), BD (n= 5) and matched controls (n=62) were included in the study (Table 1). Demographic, clinical and pharmacological data were obtained during a case history review conducted using the Diagnostic Instrument for Brain Studies (DIBS), as described previously ^90^. Tissue collection and processing was performed as described previously ^90^. Postmortem brains were collected with approval from the Ethics Committee of the Victorian Institute of Forensic Medicine and all tissue was collected by this Institute after gaining written consent of the next of kin. This study was approved by the Human Ethics Committee of Melbourne Health ^90^. All tissue was obtained from the Victorian Brain Bank at the Florey Institute for Neuroscience and Mental Health. Brodmann area 9 (BA 9) was taken from the lateral surface of the frontal lobe from an area comprising the middle frontal superior gyrus to the inferior frontal sulcus of the left hemisphere.

#### Data set 2: Human whole blood data from the Schizophrenia Exome Sequencing Meta-Analysis (SCHEMA) consortium

The SCHEMA Phase I dataset (GRCh37/hg19) containing exomes from 24,248 cases and 97,322 controls was used for analysis of rare coding variants ^46^. 244 genes have been identified to be associated with schizophrenia at P < 0.01, 231 of which are present in Dataset 1.

#### Data set 3: Human postmortem brain samples (snRNA-seq data)

Postmortem orbitofrontal cortex tissues from Brodmann Area 11 (BA11) from two neurotypical individuals (Table 2) were used for single nuclei RNA-sequencing (snRNA-seq). These brain tissues were fresh-frozen and obtained from the NSW Brain Tissue Resource Centre in Sydney, Australia. Informed consent was given by both donors or their next of kin for brain autopsy. Ethical approval for this study was obtained from the Human Research Ethics Committees at the University of Wollongong (HE2018/351) and the Ludwig-Maximilians-Universität Munich (17-085 and 18-393). BA 11 was dissected from the 3rd 8-10mm coronal slice from each fresh hemisphere for each subject.

### Gene expression data

#### Data set 1: Exon arrays

RNA preparation and expression array processing were carried out as described previously ^90^. Briefly, total RNA was isolated from ff100 mg frozen grey matter using 1.0 ml TRIzol reagent (Life Technologies, Scoresby, VIC, Australia). After homogenization and phase separation, the aqueous phase was added to an equal volume of 70% ethanol. RNA isolation was performed with RNeasy minikits (Qiagen, Cat. No. 74104, Chadstone Centre, VIC, Australia), with all samples treated with DNase using column digestion. DNA contamination was ruled out by PCR using specific primers for genomic DNA. RNA quantity and quality were analysed by spectrophotometry (NanoDrop; Thermo Fisher Scientific Australia, Scoresby, VIC, Australia) and by determining RNA Integrity Numbers (RINs) using an Agilent 2100 bioanalyzer (Agilent Technologies, Santa Clara, CA, USA) and all samples with RIN ≥ 6.00 were used for further analyses with Affymetrix Human Exon 1.0 ST v2 Arrays according to the manufacturer’s instructions (Affymetrix, Santa Clara, CA, USA). Following hybridization, chips were scanned and the fluorescent signals converted into a DAT file for quality control. Finally, cell intensity (CEL) files were generated for further analyses.

Reading the raw data CEL files and storing the 6,553,600 microarray probes was performed using the *oligo* version 1.50.0 package ^92^ in R 3.6.1. For background adjustment, quantile normalisation and summarization (using median-polish) to probeset level *oligos* Robust Multichip Average (RMA) algorithm performing all three steps at once was used. Probesets without any start or stop information or labelled as controls in the current NetAffx annotation file for the Human Exon 1.0 ST v2 Array, downloaded from the Affymetrix support website, were removed from the dataset. Cross-hybridized probesets (*number_cross_hyb_probes* ff 1), composed of RNA target sequences binding to short DNA probes that are not exactly their complement, and probesets lying on the X, Y and M chromosome were also excluded from the dataset.

*SVA* version 3.34.0 ^93,94^ package in R 3.6.1 was used for batch correction of known and hidden batches. Batch correction was conducted at probeset level before annotation and summarization to different genetic levels. We first removed the five known batches with *SVA’s ComBat* function and then applied surrogate variable (SV) analysis to remove hidden effects. One SV was found, which was the only one explaining high variance in the expression dataset (Figure S2C-E). Gene annotations were downloaded from GENCODE ^95^ release 19, which is the current version for the human hg19 (GRCh37) genome built on Ensembl version 74. Only protein-coding genes manually annotated by the Human and Vertebrate Analysis and Annotation (HAVANA) team were considered.

Summarization to gene, transcript, and exon-level was performed as follows: First, an annotation dataset was generated, where the GENCODE and the Affymetrix probeset information were merged by overlapping the locations in both datasets using the mergeByOverlaps function of the *GenomicAlignments* version 1.22.1 package in R 3.6.1. This annotation dataset was filtered for probesets, where at least one gene symbol correlated in both merged files and no multiple annotations were found. Second, for each sample the median of all expression values across all probes containing the same Ensembl ID was calculated. Multiple gene mappings were removed from all datasets. Some genes, transcripts and exons shared the exact same expression values in all samples leading to identical rows with different assigned IDs. This happens due to overlapping gene, transcript or exon locations. Instead of completely removing all identical rows, one row was kept and the different gene, transcript or exon IDs were merged into one ID. This led to expression values of 17,447 genes, 100,750 transcripts and 242,443 exons (all based on Ensembl IDs).

#### Data set 3: snRNA-seq

##### Nuclei extraction and library preparation

Nuclei were isolated using an adapted version of a previously published protocol ^96^. Briefly, frozen, dissected brain samples (50 -60 mg) were dounce-homogenised in 1 ml nuclei extraction buffer (0.32 M Sucrose, 3 mM Mg(Ac)_2_, 5 mM CaCl_2_, 0.1 mM EDTA, 10 mM TrisHCl pH 8.1, 0.1% IGEPAL CA-630, 40 U/ml RiboLock RNase-Inhibitor (ThermoScientific)) on ice. Homogenate was layered onto 1.8 ml of sucrose cushion (1.8 M Sucrose, 3 mM Mg(Ac)_2_, 10 mM TrisHCl pH 8.1) and ultra-centrifuged at 28100 rpm for 2.5 hours at 4°C. Supernatant was removed and nuclei pellet was resuspended in 100 µl resuspension buffer (1X PBS, 3 mM Mg(Ac)_2_, 5 mM CaCl_2_, 1% BSA, 40 U/ml RiboLock RNase-Inhibitor). Nuclei suspension was filtered through a cell strainer cap. Nuclei were stained with DAPI 1:1000 and counted using a hemocytometer. Libraries for snRNA-seq were prepared following the user guide of 10X Genomics (Chromium Single Cell 3’ Reagents kit v3) with a target recovery of 10,000 nuclei per sample. Libraries were pooled equimolarily and were treated with Illumina Free Adapter Blocking Reagent before sequencing on the NovaSeq 6000 System (Illumina, San Diego, California, USA).

##### Sequence Alignment, Filtering, Normalisation, Clustering and Cell type assignment

Sequence reads were demultiplexed using the sample index, aligned to a pre-mRNA reference and UMI were counted after demultiplexing of nuclei barcodes using Cell Ranger v3.1.0. Count matrices were further processed using Scanpy v1.4.4 ^97^. Count matrices of the two individuals were combined. Nuclei were filtered according to counts, minimum genes expressed and % of mitochondrial genes (Max counts > 50 000, Min counts < 1000, Min genes > 400, Mito % ≥ 10). Genes expressed in < 20 nuclei were removed. Data were normalised and log-transformed using Scran ^98^. Embeddings were created using BBKNN ^99^ and Louvain clustering ^100^ using highly variable genes was applied for clustering. One cluster was excluded from the analysis due to highly variable genes being driven by or containing several MT-genes. Cell types were assigned to clusters based on marker gene expression as follows (Nagy et al. ^101^ and Velmeshev et al. ^102^): Excitatory neurons: *SATB2, SLC17A7*, Layers: L2-4: *CUX2, THSD7A, L4-6: RORB, POU6F2, TSHZ2, RXFP1, L5-6: ETV1, KCNK2, PCP4*, Inhibitory neurons: *GAD1, GAD2*, Inhibitory neuron subtypes: In_PVALB: *PVALB*, In_SST: *SST*, In_VIP: *VIP, CALB2*, In_SV2C: *SV2C*, fibrous astrocytes (Astro_FB): high *GFAP, TNC, AQP4, GJA1*, protoplasmic astrocytes (Astro_PP): high *SLC1A2, AQP4, GJA1*, Microglia: *CD74, P2RY12, C3, CX3CR1*, Oligodendrocyte precursors (OPCs): *PCDH15, PDGFRA, OLIG1*, Oligodendrocytes (Oligo): *PLP1, MBP, MOBP, MOG*, Endothelial cells (Endo): *CLDN5, FN1, FLT1*.

### Genotype data, imputation and PRS in Dataset 1

Firstly, 100ng of genomic DNA was extracted from blocks of cerebellum from 169 subjects composed of 107 cases and 72 controls (see Table 1). Extractions were performed as previously described ^103,104^.

Genotyping of the samples was performed using Illumina Infinium Global Screening Arrays according to the manufacturer’s standard protocols. Quality control (QC) was conducted in PLINK 1.90b6.6^105^. QC steps on samples included removal of individuals with a missing rate > 2%, cryptic relatives (PI-HAT > 0.0125), an autosomal heterozygosity deviation (|F_het_| > 4 SD) and genetic outliers (distance in the ancestry components from the mean > 4 SD). QC steps on variants included removal of variants with a call rate < 98%, a MAF < 1%, and HWE test p-values ff 10^−6^. Furthermore, variants on non-autosomal chromosomes were excluded. Imputation was performed with IMPUTE2, following phasing in SHAPEIT, using the 1,000 genomes phase III reference panel. QC of imputed probabilities was conducted in QCTOOL v1.5 64-bit. Imputed SNPs were excluded if MAF < 1%, HWE test p-values ff 10^−6^, or an INFO metric < 0.6. SNP coordinates are given according to hg19 (n=9,164,462 SNPS).

PRS for all 169 individuals were calculated using PRSice-2 v2.2.11.b (14th Oct 2019) ^106^ using a P-value threshold of 0.01. We wanted to use the same P-value threshold for all calculated PRSs, and 0.01 was the best average value. As input, we used the hard-called imputed and quality controlled genotypes of the Dataset 1 as target, imputed genotypes from an independent cohort (recMDD ^107^, n=1,774 Caucasian individuals) as LD reference and eight different publicly available GWAS summary statistics as base dataset. Polygenic risk was measured for different psychiatric disorders using following GWAS summary statistics of the Psychiatric Genomics Consortium (PGC): schizophrenia ^9^, BD ^108^, MDD ^109^, autism spectrum disorder (ASD) ^110^, attention deficit/hyperactivity disorder (ADHD) ^111^ and cross disorder (CDG)^8^. We additionally generated PRS for two non-psychiatric phenotypes as negative controls: the Social Science Genetic Association Consortium (SSGAC) for educational attainment (EA) ^112^ and the DIAbetes Genetics Replication And Meta-analysis (DIAGRAM) Consortium for type 2 diabetes (T2D) ^113^ (Table S13i). Proportion of missing SNPs per GWAS summary statistic were on average 15.35% and median 15.04% (14.33% for ADHD, 14.32% for ASD, 19.46% for BD, 15.34% for CDG, 14.88% for EA, 14.07% for MDD, 15.19% for schizophrenia, 15.22% for T2D). P-value, pseudo-R^2^ based on Cox & Snell ^114^ and Cragg & Uhler (Nagelkerke) ^115^ approach were calculated using a fitted linear model approach and the Nagelkerke function of the *rcompanion* version 2.3.25 package in R (Table S13ii).

### Phenotype data in Dataset 1

For three samples, no pH value was given, so we assigned them the mean value of all pH values from all other samples. We calculated the first ten ancestry dimensions (Dim1-10) using multidimensional scaling (MDS) based on raw Hamming distances in PLINK. Samples did not group into different subpopulations, although some samples were known to be of Asian (n=5) and mixed, Asian and Caucasian (n=1), origin, while the majority (n=123) were of Caucasian descent (Figure S2A). Only the Eigenvalue of the first four genetic ancestry dimensions (Dim1-4) changed verifiably, so only these were used in further analyses (Figure S2B). Based on Stenbacka et al. ^116^, the cause of death was divided into three categories: 1) “natural” for all natural causes, e.g. ischemic heart disease or pneumonia, 2) “violent” for violent suicides or accidents, such as hanging, drowning or a car accident, and 3) “non-violent” for all types of poisoning. A visual overview of all phenotypes is given in Figure S2D-E.

### Differential expression analysis (DEA)

For the differential expression analyses, we collapsed all cases (schizophrenia, BD, MDD) and *limma* version 3.42.2 package in R 3.6.1 was used. We calculated the effect of case-control status on the difference in gene expression controlling for Age, Sex, pH, PMI, RIN, RIN^2^, Suicide and Type of Death (Natural, Violent and Non-Violent) as well as the first four dimensions of the genotype defined ancestry (Dim1-Dim4) and one found surrogate variable (SV1), which was not correlated to any other covariate of the model (Figure S2C,E). Expression matrices of 17,447 genes, 100,750 transcripts and 242,443 exons coming from 169 samples (107 cases and 62 controls) after preprocessing were used for the analysis. The differential expression analysis was performed for each genetic level separately and only significant (FDR < 0.1) hits were considered for further analysis. For transcript- and exon-level expression the FDR was calculated as follows: 1) p-values were generated using *limma*, correcting for all covariates, 2) for each Ensembl gene ID only the transcript or exon with the lowest p-value was taken generating a new expression matrix with only one transcript or exon per gene, and 3) *limma* was used to recalculate the FDR on the new expression matrix. We performed the differential expression analysis not only for diagnosis but all given covariates correcting for all other covariates of the model (Figure S2, Figure S3A and Table S1 -Table S6).

### eQTL analysis and eSNP clumping

For the eQTL analysis, all imputed SNPs with a minor allele frequency (MAF) < 5% were removed (n=6,830,577 SNPs). These SNPs were fitted to the expression of gene (n=17,447), transcript (n=100,750) and exon-level (n=242,443) using the additive linear model of MatrixEQTL^117^, adjusting for the same covariates (age, sex, pH, PMI, RIN, RIN2, suicide, ToD, Dim1-4, SV1) as in the DEA. The *cis*-eQTL output threshold was set to 0.05, the physical distance to 1Mb, no FDR memory was saved and no trans-eQTLs were calculated (threshold set to 0). For identity, the error covariance matrix was set to numeric and the minimal p-value by gene SNP was not calculated. Frequently, eQTL SNPs (eSNPs) are in LD and thus more often associated with each other than by chance. To find these linked eSNPs, we performed clumping using PLINK v1.90b6.7 64-bit (2nd Dec 2018) ^105^. For the clumping we set an r-squared LD threshold of 0.2, a significance threshold for index SNPs at 0.05, a secondary significance threshold for clumped SNPs at 1 and a physical distance threshold of 1Mb.

### Exon eQT-Score generation

To generate expression quantitative trait scores (exon eQT-Score) we used the same procedure as for the DEA replacing diagnosis with PRS, i.e., PRS was fitted to the expression using a linear model adjusting for age, sex, pH, PMI, RIN, RIN2, suicide, ToD, Dim1-4, SV1. Each PRS was used once, resulting in exon eQT-Score for the different phenotypes of the underlying 5 GWAS used for the PRS calculation.

### Enrichment analysis

#### GWAS and epigenetic enrichment analysis

We performed an enrichment analysis using public data from Ensembl Variant Effect Predictor (VEP) ^118^, the core 15-state model (ChromHMM v1.10) of chromatin in DLPFC from the Roadmap Epigenomics Project ^43^ and the GWAS summary statistics for ADHD ^111^, ASD ^110^, BD ^108^, CDG ^8^, EA ^112^, MDD ^109^, schizophrenia ^9^ and T2D ^113^. As a background dataset, we used all eQTL SNPs overlapping with the GWAS datasets. To reduce bias, we generated 11 MAF bins for each eSNP set (gene-eSNP, transcript-eSNP and exon-eSNP) and the background SNP set by using 0.05 steps from 0 to 1. We then performed an enrichment analysis for 10,000 permutations, where we calculated the overlap of randomly selected background SNPs in the size of an eSNP set and its MAF bin distributions with one of the public datasets (VEP, ChromHMM or the GWAS summary statistics). To generate an empirical p-value, we calculated how often one of the 10,000 overlaps was higher than the actual overlap of the eSNP set to the public dataset and divided it through the number of permutations. An odds ratio (OR) was calculated by the actual overlap divided by the mean of the 10,000 overlaps of the resampling (Table S12).

#### KEGG pathway and tissue enrichment analysis

For the KEGG pathway analysis of exon level differentially expressed genes or the core set of genes, we used FUMA GENE2FUNC ^41^ excluding disease and drug treatment relations. Default parameters were used in FUMA, with the exon based gene set (n=17,496) as the background list. Tissue specificity was performed on differentially expressed genes using GTEx v8 ^16^ postmortem expression Dataset. Significant tissue enrichment was obtained with Bonferroni corrected p-values (P_bon_ ff 0.05). For the KEGG pathway enrichment only significant pathways with an FDR ≤ 5% were considered.

#### Cell-type enrichment analysis

We used snRNA-seq data (Data set 3) to identify cell types enriched among the cells with the highest mean expression of the identified core genes (n=110). Core genes and the background of all tested genes (n=17,496) were mapped to the single-nuclei expression data using Ensembl gene IDs ^119^. 106 of the core genes and 14,600 of the background genes were also detected in the single-nuclei Dataset. The cell-type distribution of the 25% cells with the highest mean expression value in the core genes were compared to the same distribution taking all background genes into account. Statistical significance of the enrichment in each cell-type was evaluated using Fisher’s exact test ^120^. In addition, we conducted cell-type specific analysis using WebCSEA on snRNAseq data from BA6 (premotor) and BA10 (frontal pole) cortices (n=6 individuals)^48^ with 107 of the core genes detected. The significance was set 0.05 using the combined p-value threshold.

#### Core gene set generation and analysis

To generate the core gene set (n=110), we overlapped the found exon-eQTL genes (n=6,401) and joint exon eQT-Score genes (n=11,102) from Dataset 1 with the genes with rare variants (n=231) from Dataset 2 using Ensembl gene IDs. Filtering for the two most significant cell types (Ex_L2-4 and Ex_L4-6), a subset of 29 genes from the core gene set was generated and visualised with a heat plot using *ggplot2* version 3.3.2 package in R 3.6.1. Individual genes were examined by cell type dimension reduction plot using Uniform Manifold Approximation and Projection (UMAP) ^121^ of *Scanpy* ^97^ version 1.4.5 package in python 3.8.1, scatter plot and box plot using *ggplot2*.

## Data Availability

Expression data of dataset 1 in the present study are available online at GEO (GSE208338) and the genotypes are available upon reasonable request to the authors.
Genes altered by rare coding variants from SCHEMA exome sequencing results in dataset 2 are available online at https://schema.broadinstitute.org/downloads.
snRNA-seq data of dataset 3 in the present study are available online at GEO (GSE205642).

https://www.ncbi.nlm.nih.gov/geo/query/acc.cgi?acc=GSE205642

https://drive.google.com/drive/folders/1JRyThjCwb7JFE6LdQQiAHwXQZFIO2o0V?usp=sharing

https://www.ncbi.nlm.nih.gov/geo/query/acc.cgi?acc=GSE208338

https://schema.broadinstitute.org/downloads

## ACKNOWLEDGEMENTS

The Victorian Brain Bank tissue collection was supported by the National Health and Medical Research Council (NHMRC; Australia; grant number 566967) and the Cooperative Research Centre (CRC) for Mental Health and the Operational Infrastructure Support from the Victorian State Government. We would additionally like to thank Geoff Pavey for his efforts in curating the Australian Victorian Brain Bank. The New South Wales Brain Tissue Resource Centre at the University of Sydney was supported by the University of Sydney. Research reported in this publication was supported by the National Institute of Alcohol Abuse and Alcoholism of the National Institutes of Health under Award Number NIAAA012725-15. The content is solely the responsibility of the authors and does not represent the offcial views of the National Institutes of Health. Dr Knauer-Arloth was supported by the Brain Behaviour Research Foundation (NARSAD Young Investigator Grant, #28063). Dr Matosin was supported by an Al and Val Rosenstrauss Fellowship from the Rebecca L Cooper Medical Research Foundation and grants from the Brain Behaviour Research Foundation (NARSAD Young Investigator Grant, #26486) and the Rebecca L. Cooper Medical Research Foundation (#PG2020645). We thank Elisabeth B. Binder for the useful discussions during the project implementation and Vanessa Schmoll for her support with genotypes of Dataset 1. We thank Miriam Gagliardi and Simone Röh for assistance with dataset 3. We thank Ghalia Rehawi for feedback on the enrichment analysis. We thank all participants for their contribution to this study.

## CONFLICT OF INTEREST

All authors do not have conflicts to declare.

## DATA AND CODE AVAILABILITY

Code for the analysis is available at GitHub: https://github.com/cellmapslab/PostmortemBrainAnalysis. Expression data of Dataset 1 are available on GEO (GSE208338) and the genotypes are available upon approved request. snRNA-seq data of Dataset 3 are available on GEO (GSE205642). The web links for the publicly available datasets used in the study are as follows: SCHEMA results to integrate rare coding variants: https://schema.broadinstitute.org/downloads, 15-core state model of chromatin from Roadmap Epigenomics Roadmap (ChromHMM v1.10) for functional annotation of eSNPs: https://egg2.wustl.edu/roadmap/web_portal/chr_state_learning.html#core_15state, CommonMind Consortium for result comparison: http://CommonMind.org,> Current NetAffx probeset annotation file for the Affymetrix HuEx 1.0 ST v2 microarray: http://www.affymetrix.com/Auth/analysis/downloads/na36/wtexon/HuEx-1_0-st-v2.na36.hg19.probeset.csv.zip, DIAbetes Genetics Replication And Meta-analysis (DIAGRAM) Consortium for the type 2 diabetes (T2D) GWAS summary statistic: https://diagram-consortium.org/downloads.html, GENCODE release 19 (GRCh37.p13) annotation file: http://ftp.ebi.ac.uk/pub/databases/gencode/Gencode_human/release_19/gencode.v19.annotation.gtf.gz, Genotype-Tissue Expression (GTEx) V8 for annotation/enrichment analysis and result comparison: https://www.gtexportal.org/home/datasets, Human Ageing Genomic Resources (HAGR) for known human ageing genes: https://genomics.senescence.info/genes/human_genes.zip, National Center for Biotechnology Information (NCBI) for GRCh37/hg19 reference genome of the index-patient rare variant dataset: http://www.ncbi.nlm.nih.gov/assembly/2758/, Psychiatric Genomics Consortium (PGC) for GWAS summary statistics of psychiatric disorders: https://www.med.unc.edu/pgc/, Social Science Genetic Association Consortium (SSGAC) for the educational attainment (EA) GWAS summary statistic: https://www.thessgac.org/data, Web-based Cell-type-Specific Enrichment Analysis (WebCSEA): https://bioinfo.uth.edu/webcsea/.

## AUTHOR CONTRIBUTION STATEMENTS

K.W. performed main computational analysis, data visualizations and wrote the first draft of the manuscript. N.M. provided intellectual input and performed critical revisions of the manuscript, led the collaboration for exon array resources (dataset 1), and carried out the design and tissue acquisition for snRNA-seq analysis (Dataset 3). J.K.-A. and N.S.M. (Lead Contacts) jointly conceived, designed and supervised the study. K.W., J.K.-A. and N.S.M. equally contributed to the analysis strategy design, result interpretation and writing the manuscript. A.S.F. performed snRNA-seq library preparation and conducted snRNA-seq analysis with N.G. (Dataset 3). M.R, F.D., H.T. and A.K. were involved in rare variants analysis. B.D. was involved in the collection and selection of appropriate postmortem tissue samples (Dataset 1). N.S.M., J.K.-A., N.M., B.D., F.D. and F.J.T. acquired funding for the study. All other authors saw, had the opportunity to comment on, and approved the final manuscript.

## Notes

### Competing Interest Statement

The authors have declared no competing interest.

### Clinical Protocols

https://www.ncbi.nlm.nih.gov/pmc/articles/PMC3233860/pdf/jove-20-914.pdf

### Author Declarations

Ethics committee/IRB of the Victorian Institute of Forensic Medicine gave ethical approval for this work. Ethics committee/IRB of the Human Ethics Committee of Melbourne Health gave ethical approval for this work. Ethics committee/IRB of the Central Institute of Mental Health Mannheim gave ethical approval for this work. Ethics committee/IRB of the University Hospital Bonn gave ethical approval for this work. Ethics committee/IRB of the Human Research Ethics Committees at the University of Wollongong gave ethical approval for this work. Ethics committee/IRB of the Ludwig-Maximilians-Universitaet Muenchen gave ethical approval for this work.

### Summary of Updates

Dataset 2, which contains rare variant altered genes, was replaced with the SCHEMA exome sequencing results since they used a much larger cohort and the results are publicly available. Polygenic risk scores (PRS) were recalculated for the most recent genome-wide association study (GWAS) summary statistics for bipolar disorder and schizophrenia, and eQT scores and the core gene set were updated accordingly. All figures, tables, and text have been adjusted to reflect the new results.

