## Supplementary methods figures and table legends for "Variant-risk-exon interplay impacts circadian rhythm and dopamine signaling pathway in severe psychiatric disorders"

### 1. Study samples, tissue collection and processing

#### *Dataset 1: Human post-mortem brain samples (exon array data)*

The primary cohort (dataset 1) has been previously described in Scarr et al. [1] and Dean et al. [2]. Briefly, postmortem DLPFC tissue from 169 adult subjects aged 18-87 years with either SCZ (n = 68), MDD (n = 24), BD (n = 15) and matched controls (n = 62) were included in the study (Table 1a). Demographic, clinical and pharmacological data were obtained during a case history review conducted using the Diagnostic Instrument for Brain Studies (DIBS), as described previously [1]. Tissue collection and processing was performed as described previously [1]. Postmortem brains were collected with approval from the Ethics Committee of the Victorian Institute of Forensic Medicine and all tissue was collected by this Institute after gaining written consent of the next of kin. This study was approved by the Human Ethics Committee of Melbourne Health [1]. All tissue was obtained from the Victorian Brain Bank at the Florey Institute for Neuroscience and Mental Health. Brodmann area 9 (BA 9) was taken from the lateral surface of the frontal lobe from an area comprising the middle frontal superior gyrus to the inferior frontal sulcus of the left hemisphere.

PRS for all 169 individuals were calculated using PRSice-2 v2.2.11.b (14th Oct 2019) [17] using a P-value threshold of 0.01. As input, we used the hard-called imputed and quality controlled genotypes of Dataset 1 as target, imputed genotypes from an independent cohort (recMDD [18], n = 1,774 Caucasian individuals) as LD reference and eight different publicly available GWAS summary statistics as base dataset. Polygenic risk was measured for different psychiatric disorders using following GWAS summary statistics of the Psychiatric Genomics Consortium (PGC): attention deficit/hyperactivity disorder (ADHD) [19], autism spectrum disorder (ASD) [20], bipolar disorder (BD) [21], cross disorder (CDG) [22], major depressive disorder (MDD) [23] and schizophrenia (SCZ) [24]. We additionally generated PRS for two non-psychiatric phenotypes as negative controls: the Social Science Genetic Association Consortium (SSGAC) for educational attainment (EA) [25] and the DIAbetes Genetics Replication And Meta-analysis (DIAGRAM) Consortium for type 2 diabetes (T2D) [26] (Table S13i). P-value, pseudo-$R^2$ based on Cox & Snell [27] and Cragg & Uhler (Nagelkerke) [28] approach were calculated using a fitted linear model approach and the Nagelkerke function of the *rcompanion* version 2.3.25 package in R (Table S13ii).

### Supplementary Figures and Legends

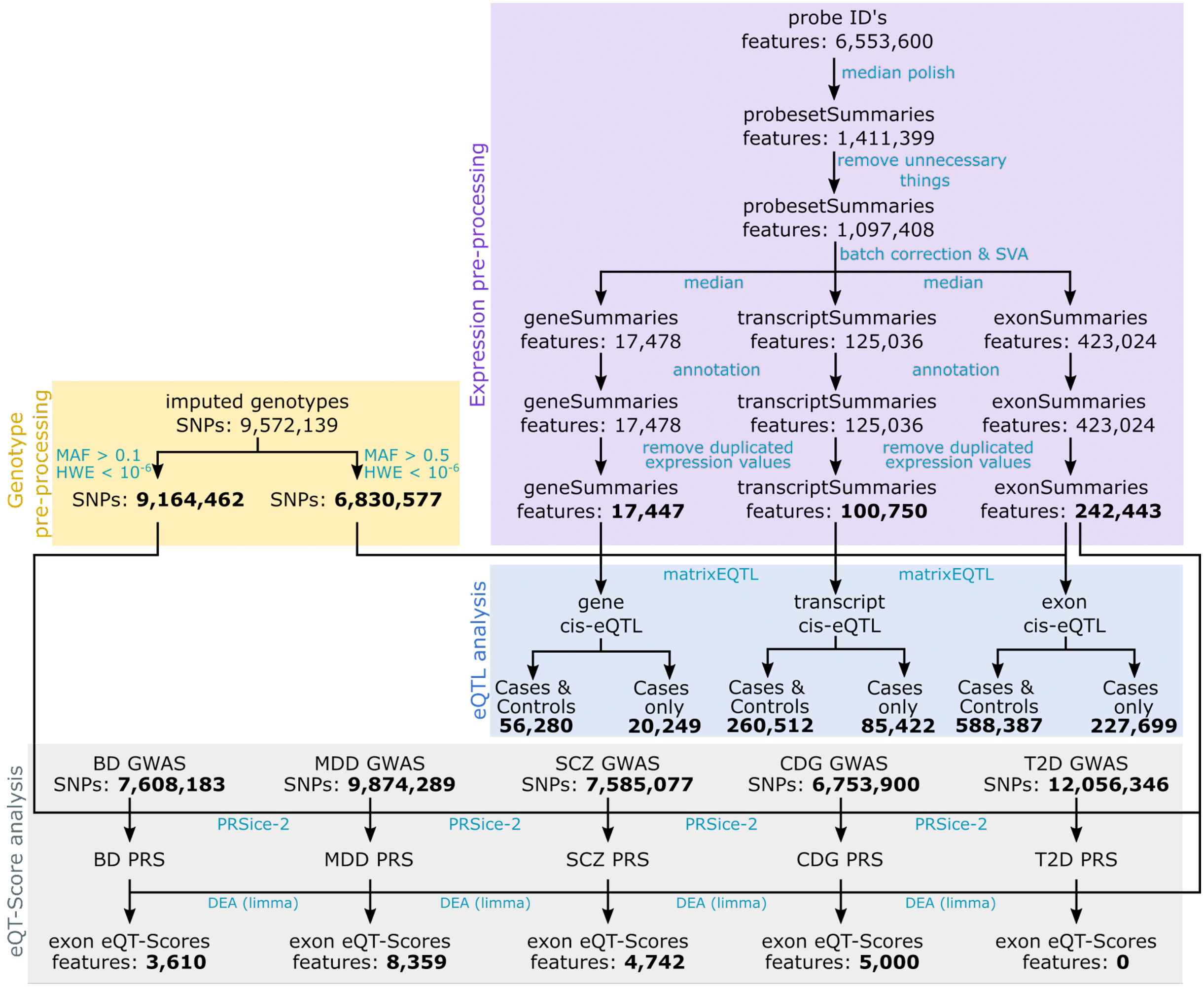


**Figure S1: Overview of the microarray preprocessing steps.** The genotype preprocessing (yellow box) splits into two sections: 1) The genotypes are filtered for a minor allele frequency (MAF) > 0.1 and a Hardy-Weinberg equilibrium (HWE) < $10^{-6}$. Resulting SNPs were used for polygenic risk score (PRS) calculations and eQT-Score analysis (light grey box). 2) Genotypes were filtered for MAF < 0.05 and HWE< $10^{-6}$ for the eQTL analysis. The expression preprocessing (purple box) comprises several steps, including a self-made median summarization to gene, transcript and exon-level summarization. Both genotypes and expressions were combined for the eQTL analysis (blue box). The turquoise texts next to the arrows show either the used method/tool or a short description of the task done at this step. Bold numbers display the number of resulting data values used for the following steps.

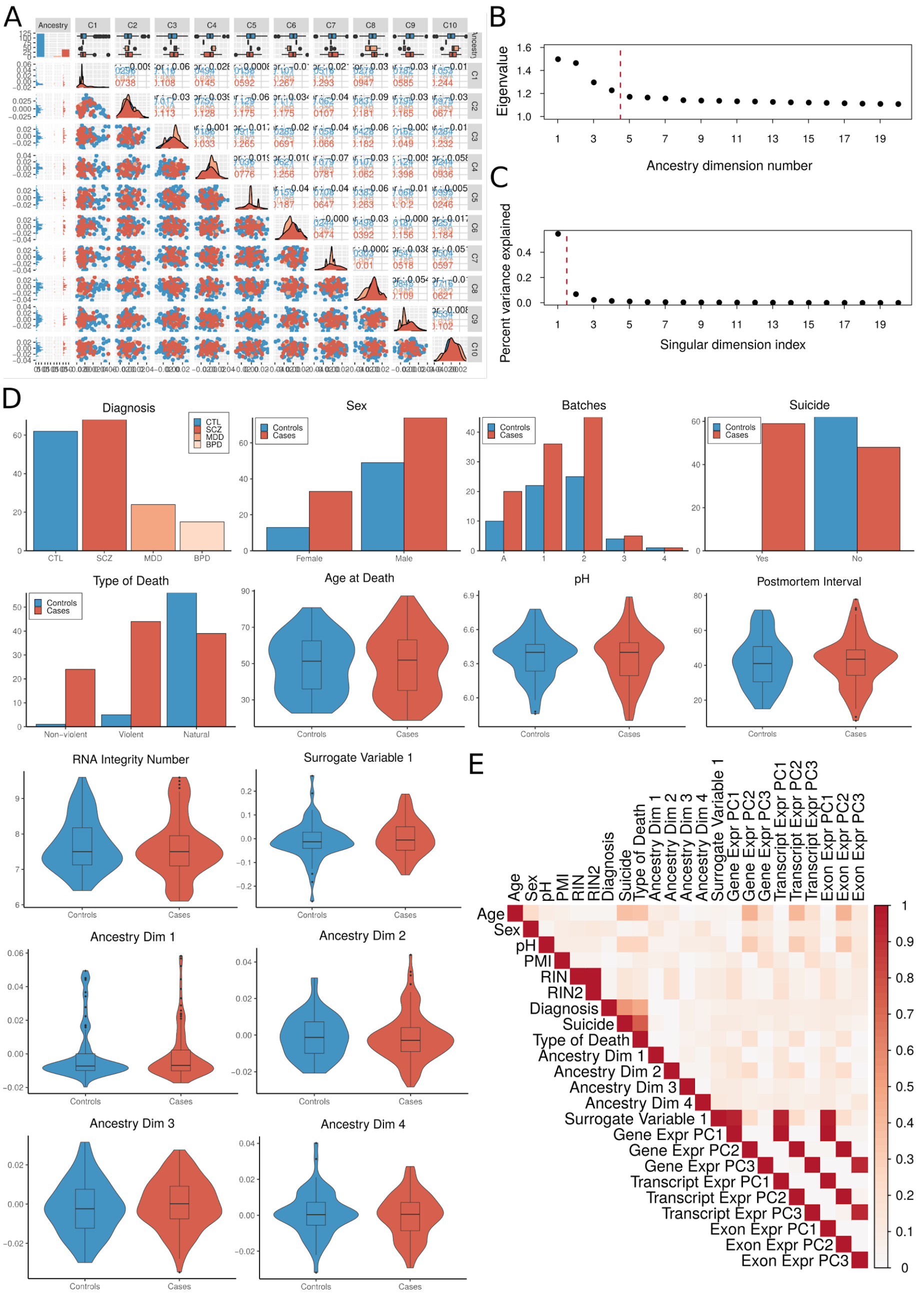


**Figure S2: Visual presentation of covariate distributions.** A) MDS plot comparing the known ancestry to the first two genetic ancestry dimensions. The first plot in the upper left corner shows a barplot of the known ancestry counts. In the column underneath facet histograms and in the first row boxplots show the combination of both datasets. The rest of the plot displays in the upper triangle the Pearson correlation coefficient, in the diagonal the distributions and in the lower triangle PCA plots of the different dimensions to each other. No visible clusters were formed anywhere. B) Eigenvalues of the ancestry dimensions, which are calculated from the available genotype data of the samples. The threshold (red dotted line) represents the point at which the eigenvalues do not vary greatly anymore. Resulting in the first four ancestry dimensions to correct for. C) Percentage of explained variance shown for the first twenty surrogate variables, which are calculated with the SVA package in R. The red dotted line shows the threshold, which indicates that the first surrogate variable (SV1) should be embedded in the model. D) Distribution shown for cases (red) vs. controls (blue) for diagnosis, microarray batches and all covariates included in the linear model. Categorical data is presented as barplots and continuous data as violin plots including smaller boxplots inside. E) Correlation plot of all covariates and the first three principal components for the gene, transcript and exon expression.

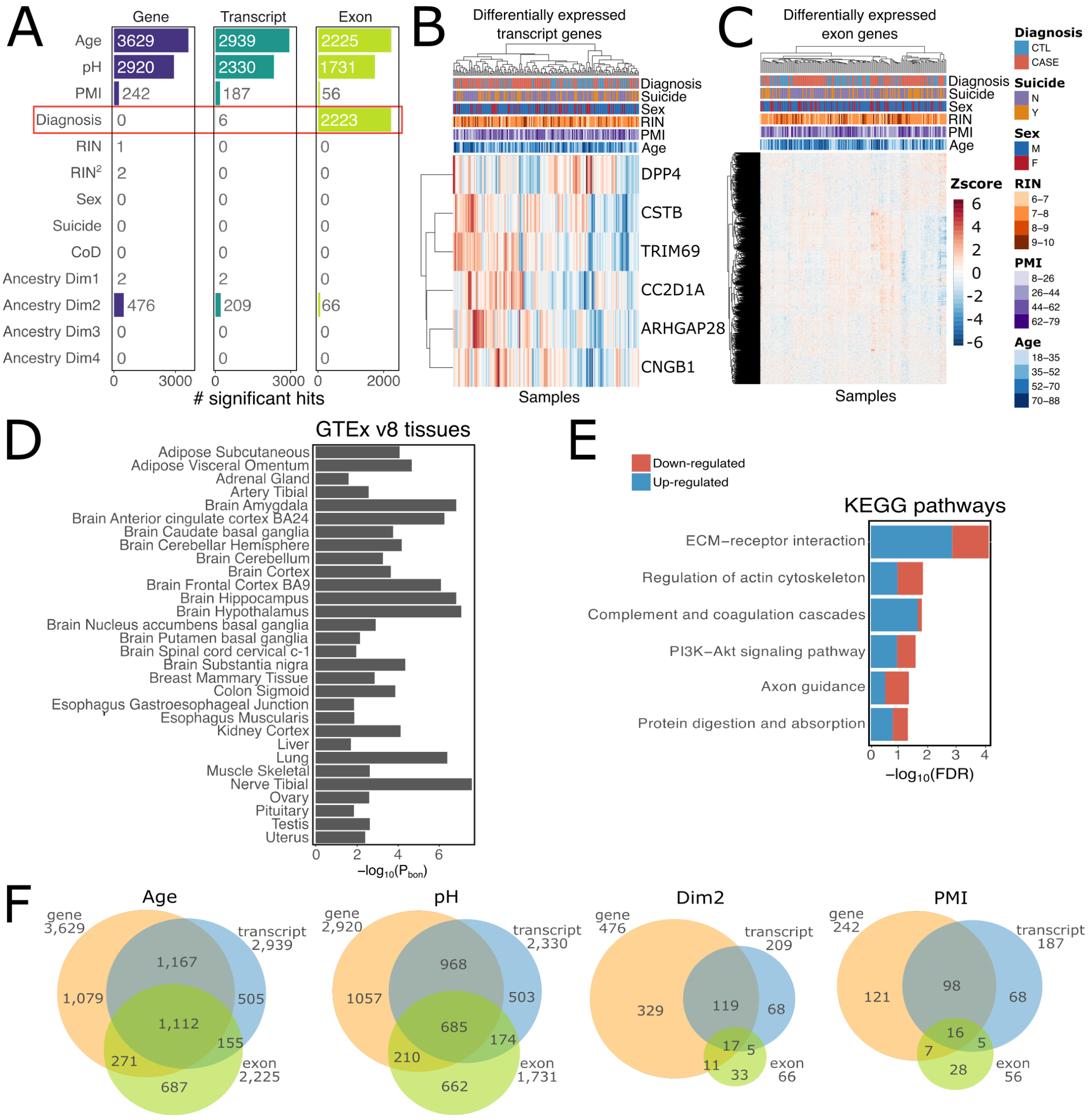


**Figure S3: Differential expression results.** A) Bar plots showing the differentially expressed gene hits (FDR < 0.1) for all model covariates on gene (orange), transcript (blue) and exon (green) level. The red box highlights the findings between cross-disorder cases and controls for diagnosis. B) Heatmaps of the six transcript-level differentially expressed genes and C) 2,223 exon-level differentially expressed genes. Heatmaps with default Euclidean distance and complete clustering method was used. D) Tissue enrichment of the 2,223 exon-level differentially expressed genes using FUMA [31] with GTEx v8 [32] expression data. Only significantly enriched (Bonferroni corrected p-value; $P_{bon}$ ≥ 0.05) tissues are shown. E) Pathway enrichment of the 2,223 exon-level differentially expressed genes using FUMA with KEGG [33], where disease and drug development pathways were excluded. Only significantly enriched (FDR < 0.05) KEGG pathways are shown colored with the proportion of up- and down-regulated genes. F) Venn diagrams showing the overlap of differentially expressed gene hits at gene, transcript and exon-level for Age, pH, genetic ancestry dimension 2 (Dim2) and postmortem interval (PMI).

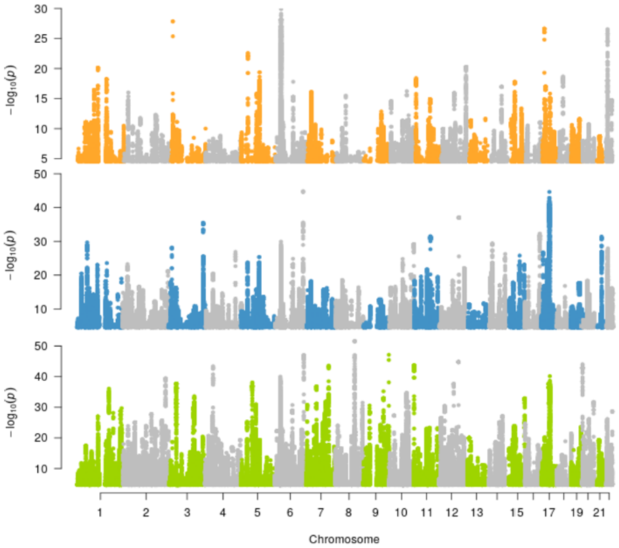


**Figure S4: eQTL results.** Manhattan plots of gene, transcript and exon eQTLs.

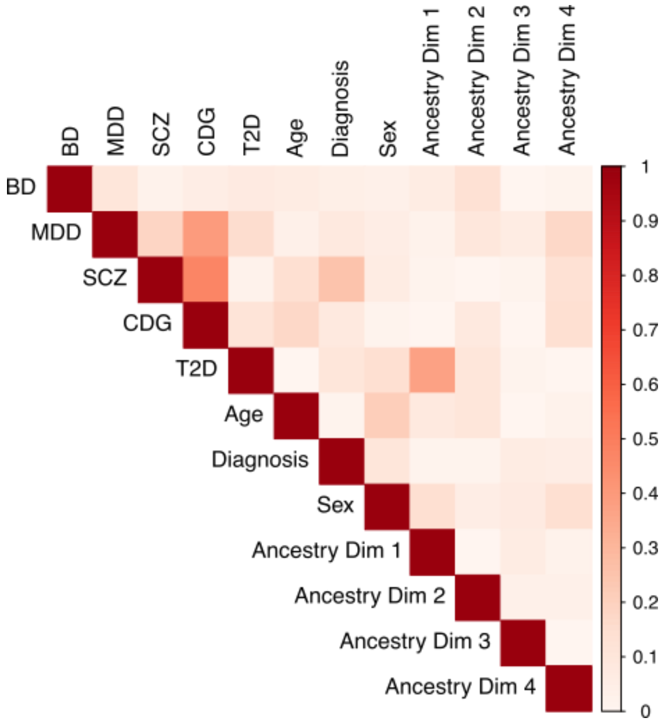


**Figure S5: Polygenic risk score (PRS) results.** Corrplot showing the correlations between diagnosis, age, sex, the four ancestry dimension covariates and the polygenic risk scores (PRS). PRS were generated with PRSice-2 and following GWAS summary statistics: from the PGC for BD (2021), CDG (2019), MDD (2019) and SCZ (2022) and one negative control from DIAGRAM for type 2 diabetes (T2D, 2017). Correlations between PRS and the covariates were calculated in R 4.2.1 with the canCorPairs function of the variancePartition version 1.26.0 package, which uses a Canonical Correlation Analysis (CCA) assessing the degree to which two vectors covary and contain the same information. No high correlation between PRS and the given covariates was found.

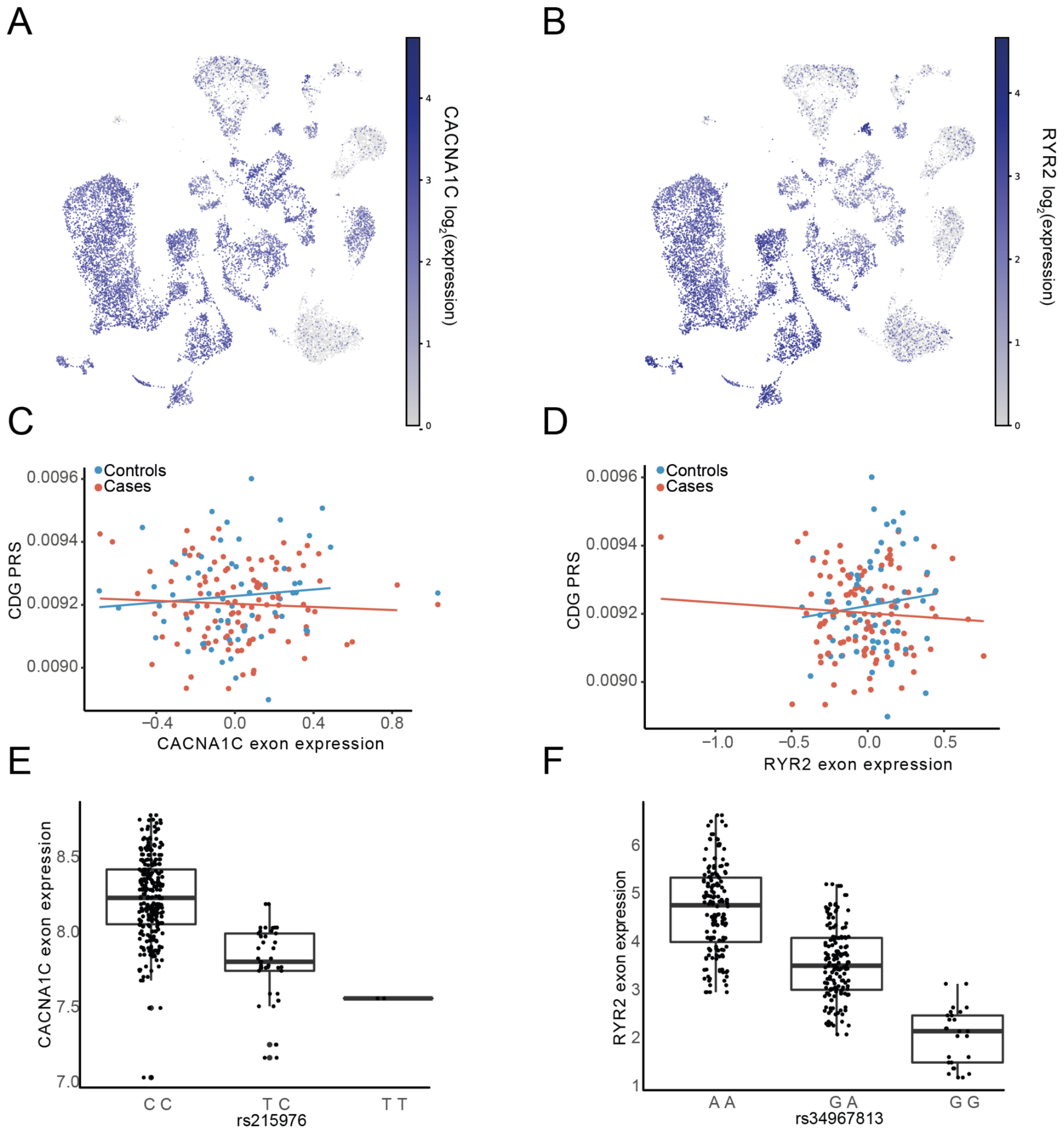


**Figure S6: *CACNA1C* and *RYR2* gene results.** A-B) Umap of the A) Calcium Voltage-Gated Channel Subunit Alpha1 C (*CACNA1C*) expression, and B) Ryanodine Receptor 2 (*RYR2*) expression, where grey denotes minimal expression and blue high expression. C-D) Scatter plots showing the cross-disorder (CDG) polygenic risk score (PRS) against the C) ENSE00001624188.1 exon expression of *CACNA1C*, and D) ENSE00001165690.1 exon expression of *RYR2*. E-F) Boxplots of the top exon-eQTL of E) *CACNA1C*: ENSE00003573219.1 expression for SNP rs215976, and F) *RYR2*: ENSE00001165512.1 expression for SNP rs34967813.

**Supplementary Tables**

**Table S1**: i) Gene-, ii) transcript- and ii) exon-level differentially expressed genes for diagnosis. Note. Ensembl Gene ID = stable Ensembl gene identifier; Ensembl Transcript ID = stable Ensembl transcript identifier; Ensembl Exon ID = stable Ensembl exon identifier; Location = gene, transcript or exon location including chromosome, start and end position; Gene Symbol = HGNC or MGI-approved gene symbols; logFC = estimate of the log2-fold-change corresponding to the effect or contrast; Average Expression = average log2-expression over all samples; T-statistic = moderated t-statistic (log2FC divided by its standard error); P-value = raw p-value; FDR estimate = Benjamini-Hochberg false discovery rate adjusted p-value; Log-odds Ratio = log-odds that gene, transcript or exon is differentially expressed.
**Table S2:** i) Gene-, ii) transcript- and iii) exon-level differentially regulated genes for age at death. Same column labels as S1.
**Table S3:** i) Gene-, ii) transcript- and iii) exon-level differentially regulated genes for pH. Same column labels as S1.
**Table S4:** i) Gene-, ii) transcript- and iii) exon-level differentially regulated genes for the first genetic ancestry dimension (PC1). Same column labels as S1.
**Table S5:** i) Gene-, ii) transcript- and iii) exon-level differentially regulated genes for the second genetic ancestry dimension (PC2). Same column labels as S1.
**Table S6:** i) Gene-, ii) transcript- and iii) exon-level differentially regulated genes for postmortem interval (PMI). Same column labels as S1.
**Table S7:** i) Overlap of differentially expressed genes for age with the Human Ageing Genomic Resources (HAGR) database. ii) BIC score for each covariate. iii) BIC scores for the quadratic terms of quasi continuous covariates.
**Table S8**: List of gene-level cis-eQTL results (geQTLs). Note. SNP = dbSNP rs ID; Ensembl Gene ID = stable Ensembl gene identifier; Gene Symbol = HGNC or MGI-approved gene symbols; Beta = effect size estimate; T-statistic = test statistic (t-test); P-value = raw p-value; FDR estimate = Benjamini-Hochberg false discovery rate adjusted p-value; SNP Location = SNP chromosome and base pair position; SNP_allele1 = reference allele of the SNP; SNP_allele2 = alternate allele of SNP; Gene Location = gene location given as chromosome, start and end position; Gene Strand = DNA strand where the gene is located; eQTL Type = information if eQTL is of type cis or trans.
**Table S9**: List of transcript-level cis-eQTL results. Same column labels as S7 + Transcript ID = stable Ensembl transcript identifier; Transcript location = position of transcript with chromosome, start and end position.
**Table S10**: List of exon-eQTL results. Same column labels as S9 + Exon ID = stable Ensembl exon identifier; Exon Location = position of exon with chromosome, start and end position.
**Table S11:** i) Counts of eQTL hits (FDR < 5%). ii) Counts of clumped eQTL hits (FDR < 5%).
**Table S12:** i) eSNP annotation to Variant Effect Predictor (VEP) categories. ii) eSNP annotation to ChromHMM DLPFC categories. iii) eSNP annotation to genome-wide association studies (GWAS) of different phenotypes. iv) eSNP annotation to CMC eQTL and isoQTL SNPs. v) eSNP annotation to GTEx DLPFC BA9 eQTL and sQTL SNPs.
**Table S13:** i) GWAS summary statistics overview used for the PRS calculation. ii) P-values, Nagelkerke and Cox & Snell $R^2$ of PRSice-2 calculated PRS. Values are given for the p-value thresholds 1, 0.01, 5e-08 and the best PRSice-2 calculated threshold.
**Table S14:** List of unique Ensembl Gene IDs and gene symbols from the BD, MDD and SCZ joint exon eQT-Score dataset.
**Table S15:** List of unique Ensembl Gene IDs and gene symbols from the overlapping core set (n = 110) of exon-eQTL genes, genes from rare coding variants from SCHEMA consortium and the joint exon eQT-Score dataset.
